# AI-based volumetric six-tissue body composition quantification from CT cardiac attenuation scans enhances mortality prediction: multicenter study

**DOI:** 10.1101/2024.07.30.24311224

**Authors:** Jirong Yi, Anna M. Michalowska, Aakash Shanbhag, Robert J.H. Miller, Jolien Geers, Wenhao Zhang, Aditya Killekar, Nipun Manral, Mark Lemley, Mikolaj Buchwald, Jacek Kwiecinski, Jianhang Zhou, Paul B. Kavanagh, Joanna X. Liang, Valerie Builoff, Terrence D. Ruddy, Andrew J. Einstein, Attila Feher, Edward J. Miller, Albert J. Sinusas, Daniel S. Berman, Damini Dey, Piotr J. Slomka

## Abstract

**Background:** Computed tomography attenuation correction (CTAC) scans are routinely obtained during cardiac perfusion imaging, but currently only utilized for attenuation correction and visual calcium estimation. We aimed to develop a novel artificial intelligence (AI)-based approach to obtain volumetric measurements of chest body composition from CTAC scans and evaluate these measures for all-cause mortality (ACM) risk stratification.

**Methods:** We applied AI-based segmentation and image-processing techniques on CTAC scans from a large international image-based registry (four sites), to define chest rib cage and multiple tissues. Volumetric measures of bone, skeletal muscle (SM), subcutaneous, intramuscular (IMAT), visceral (VAT), and epicardial (EAT) adipose tissues were quantified between automatically-identified T5 and T11 vertebrae. The independent prognostic value of volumetric attenuation, and indexed volumes were evaluated for predicting ACM, adjusting for established risk factors and 18 other body compositions measures via Cox regression models and Kaplan-Meier curves.

**Findings:** End-to-end processing time was <2 minutes/scan with no user interaction. Of 9918 patients studied, 5451(55%) were male. During median 2.5 years follow-up, 610 (6.2%) patients died. High VAT, EAT and IMAT attenuation were associated with increased ACM risk (adjusted hazard ratio (HR) [95% confidence interval] for VAT: 2.39 [1.92, 2.96], p<0.0001; EAT: 1.55 [1.26, 1.90], p<0.0001; IMAT: 1.30 [1.06, 1.60], p=0.0124). Patients with high bone attenuation were at lower risk of death as compared to subjects with lower bone attenuation (adjusted HR 0.77 [0.62, 0.95], p=0.0159). Likewise, high SM volume index was associated with a lower risk of death (adjusted HR 0.56 [0.44, 0.71], p<0.0001).

**Interpretations:** CTAC scans obtained routinely during cardiac perfusion imaging contain important volumetric body composition biomarkers which can be automatically measured and offer important additional prognostic value.

**Research in context:** *Evidence before this study:* Fully automated volumetric body composition analysis of chest computed tomography attenuation correction (CTAC) can be obtained in patients undergoing myocardial perfusion imaging. This new information has potential to significantly improve risk stratification and patient management. However, the CTAC scans have not been utilized for body composition analysis to date. We searched PubMed and Google Scholar for existing body composition related literature on June 5, 2024, using the search term (“mortality”) AND (“risk stratification” OR “survival analysis” OR “prognostic prediction” OR “prognosis”) AND (“body composition quantification” OR “body composition analysis” OR “body composition segmentation”). We identified 34 articles either exploring body composition segmentation or evaluating clinical value of body composition quantification. However, to date, all the prognostic evaluation is performed for quantification of three or fewer types of body composition tissues. Within the prognostic studies, only one used chest CT scans but utilized only a few specified slices selected from the scans, and not a standardized volumetric analysis. None of these previous efforts utilized CTAC scans, and none included epicardial adipose tissue in comprehensive body composition analysis.

*Added value of this study:* In this international multi-center study, we demonstrate a novel artificial intelligence-based annotation-free approach for segmenting six key body composition tissues (bone, skeletal muscle, subcutaneous adipose tissue, intramuscular adipose tissue, epicardial adipose tissue, and visceral adipose tissue) from low-dose ungated CTAC scans, by exploiting existing CT segmentation models and image processing techniques. We evaluate the prognostic value of metrics derived from volumetric quantification of CTAC scans obtained during cardiac imaging, for all-cause mortality prediction in a large cohort of patients. We reveal strong and independent associations between several volumetric body composition metrics and all-cause mortality after adjusting for existing clinical factors, and available cardiac perfusion and atherosclerosis biomarkers.

*Implications of all the available evidence:* The comprehensive body composition analysis can be routinely performed, at the point of care, for all cardiac perfusion scans utilizing CTAC. Automatically-obtained volumetric body composition quantification metrics provide added value over existing risk factors, using already-obtained scans to significantly improve the risk stratification of patients and clinical decision-making.

## Introduction

Computed tomography attenuation correction (CTAC) scans are routinely obtained in single-photon emission computed tomography (SPECT)/computed tomography (CT) cardiac perfusion imaging. However, these auxiliary scans are currently utilized only for attenuation correction and visual calcium estimation.^1^ Over 6 million patients in the United States undergo SPECT cardiac perfusion imaging annually.^2^ Expanding the scope of these scans can potentially enhance the utility of this widely used cardiovascular imaging modality.

Body composition analysis, which examines the amount and distribution of body tissues, such as skeletal muscle and adipose tissue, provides important anatomic information, especially when abnormalities are present. Sarcopenia, cachexia, and obesity are well-known to be associated with cardiovascular,^2^ oncological,^3^ and metabolic disorders.^4^ However, comprehensive body composition biomarkers have not been evaluated from cardiac perfusion CTAC scans to date. A fully automated approach to extract these measures during routine cardiac imaging could significantly enhance the actionable information available to clinicians.

While opportunistic, artificial intelligence (AI)-assisted evaluation of body composition on cross-sectional imaging modalities has gained interest recently, data on CTAC-based body composition analysis remains scarce.^2, 5–7^ Studies have shown that automatic epicardial adipose tissue (EAT) derived from CTAC can provide additional information and improve mortality prediction.^8^ Existing deep learning-based approaches for body composition segmentation have been developed for pre-selected abdominal CT slice,^2, 7, 9^ and only one study so far utilized selected three slices from CT.^5^ Until now, the full predictive value of CTAC in cardiometabolic screening remains unexplored.

We aimed to evaluate CTAC’s value for patient risk assessment, by developing a fully automated and standardized AI-based method for CTAC-derived volumetric measurements of body composition compartments including subcutaneous adipose tissue (SAT), visceral adipose tissue (VAT), intramuscular adipose tissue (IMAT), skeletal muscle (SM) and bone, together with our previously validated EAT segmentation model to predict all-cause mortality (ACM).

## Methods

### Study population

This is a retrospective study of 11035 patients who underwent SPECT/CT cardiac perfusion imaging from four different sites (Yale University, University of Calgary, Columbia University, University of Ottawa) participating in the REgistry of Fast Myocardial Perfusion Imaging with NExt generation SPECT (REFINE SPECT).^10^ The study protocol complied with the Declaration of Helsinki and approval was obtained from the institutional review boards (IRB) at each participating institute. The overall study was approved by the IRB at Cedars-Sinai Medical Center, Los Angeles, California. Baseline demographic and clinical data were obtained from the REFINE SPECT registry.^10^ The endpoint was ACM; for sites in the United States this was determined using the National Death Index, while in Canada, administrative databases were used.

### CTAC image acquisition parameters

All CTAC scans were low-dose, non-contrast-enhanced and non-electrocardiographically-gated. The scans were acquired with different scanners (GE Discovery NM/CT 570c, Philips Precedence 16P, and Siemens Symbia Intevo 16). The slice thickness range was 2.5-5.0 mm, tube current 16-30 mAs, and tube voltage 120-130 kVp. At two sites (Yale University and Columbia University) scans were performed free breathing, while at the remaining two (University of Calgary, and University of Ottawa), scans were performed using an end-expiratory breath hold.

### Clinical data and existing risk factors

Risk factors from clinical data or cardiac perfusion analysis included in the prognostic analysis included sex, age, body mass index (BMI), hypertension, diabetes mellitus, dyslipidemia, family history of coronary artery disease (CAD), smoking, stress total perfusion deficit (TPD), left ventricle ejection fraction (LVEF), and CAC score. Dedicated software (Quantitative Perfusion SPECT [QPS], Cedars-Sinai Medical Center, Los Angeles) was used to quantify automatically stress TPD and LVEF from myocardial perfusion imaging (MPI) scans at the core laboratory (Cedars-Sinai Medical Center, Los Angeles).^11^ Stress TPD <5% was considered as normal myocardial perfusion, whereas impaired LVEF was defined as LVEF <50%.^8^ CAC was segmented and quantified automatically from CTACs with a previously validated deep learning model.^12^

### Body composition segmentation and quantification

Figure 1 summarizes the study design. An open-source foundational model (TotalSegmentator) was used to segment 117 structures from CTAC scans.^13^ To reduce the effects of variations in scanned body regions due to different CTAC scan lengths, we computed the CTAC-volumetric measurements between T5 and T11 vertebrae which was covered by 96.4% of scans (Supplementary Figure 1). T5 and T11 were identified using results from TotalSegmentator.^13^ Based on segmented structures and image processing techniques, a chest rib cage was created to automatically separate SM, SAT, IMAT, and VAT inside and outside the rib cage (see Supplementary Figure 2). Hounsfield unit (HU) filtering with tissue-specific thresholds (for skeletal muscles between -29 and +150 HU, for adipose tissue between -190 and -30 HU). EAT was segmented separately from VAT by using our previously developed deep learning model.^8^ The EAT model was trained and validated using 500 CTACs from Yale University which were excluded from the analysis. Bone tissue was analyzed based on Total Segmentator segmentation and thresholding between +151 and +1200 HU.^13^ See more details in Supplementary Methods.

**Figure 1.**
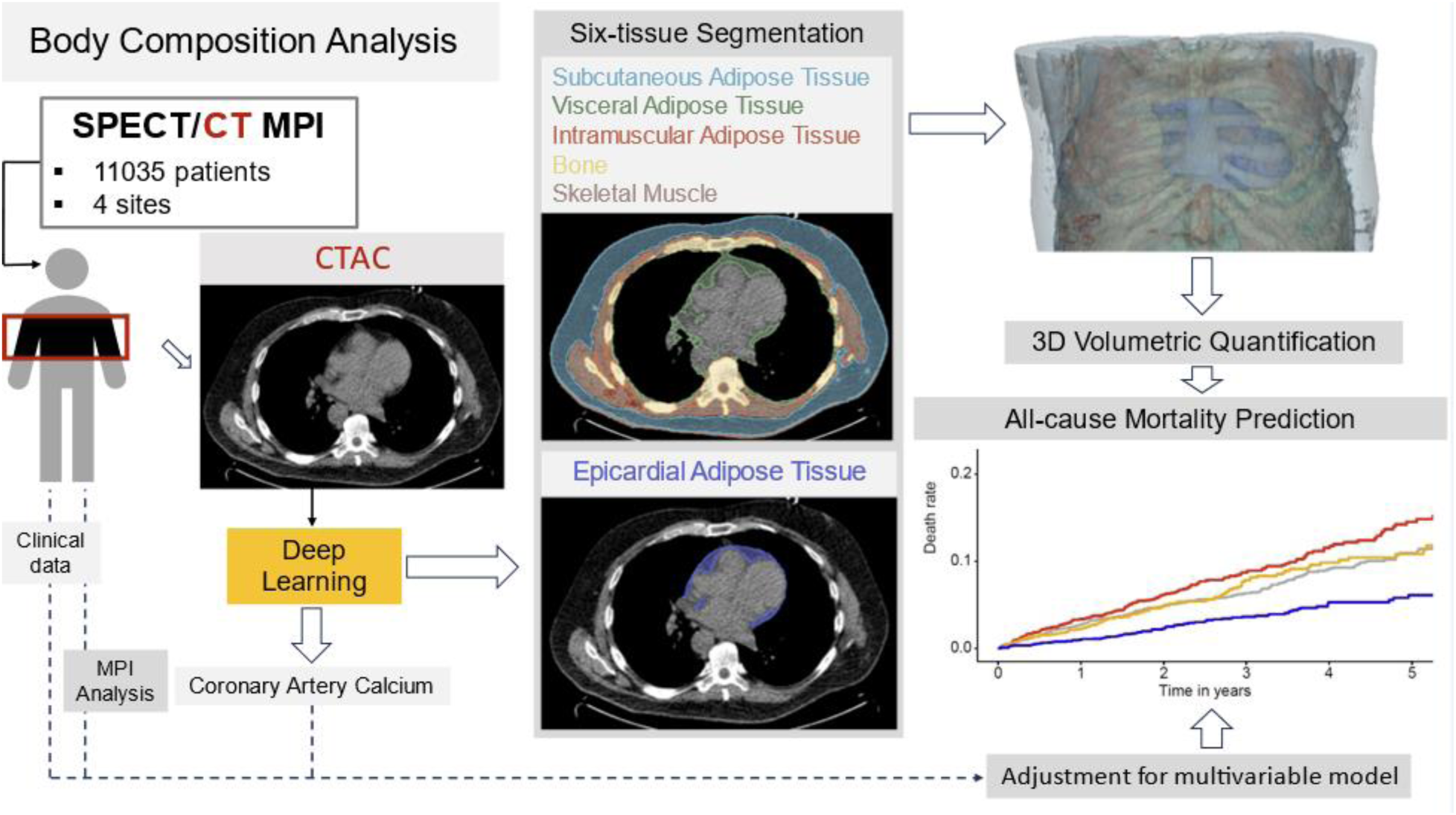
Study design: artificial intelligence-derived computed tomography attenuation correction (CTAC)-based body composition analysis. We integrated fully automated segmentation of skeletal muscles, bone, subcutaneous, intramuscular, and visceral adipose tissues with our previously validated deep-learning model for epicardial adipose tissue segmentation to predict all-cause mortality in patients undergoing myocardial perfusion imaging (MPI). SPECT – single-photon emission computed tomography.

We evaluated the prognostic value of three types of volumetric measures of six segmented body tissues including attenuation (mean of HU), standard deviation (standard deviation of the HU), volume index (volume divided by height squared, in units of cm^3^/m^2^). The ratio of skeletal muscle volume to total volume of adipose tissues (SAT, IMAT, EAT, and VAT) was also calculated. Unadjusted and multivariable models (adjusted for 11 clinical and imaging factors and the other 18 body composition quantifications) were used to evaluate associations with ACM. Prognostic evaluations were performed for the whole study group as well as for subgroups, distinguished based on race (Black or African American, White), gender (male, female), age (<65, ≥65 years), and BMI (BMI <30 kg/m^2^, BMI ≥30 kg/m^2^). Due to insufficient data for other races, we included only Black and White races in the race-based subgroup analysis.

### Statistical analysis

Continuous variables were summarized as median (interquartile range [IQR], (IQ1, IQ3)), and discrete variables were summarized as frequency (%). The Wilcoxon rank sum test was used for comparisons of continuous variables. The Pearson’s chi-squared test was used to compare categorical variables with two levels (binary variables) while the Fisher’s exact test was used for categorical variables with multiple levels. The log-rank test and the Wald test were used to evaluate statistical significance for Kaplan-Meier curves and HRs, respectively. The Youden index was used to find their optimal cutoffs for risk stratification, and hazard ratios (HRs) using Cox regression model was calculated. All statistical tests were two-sided, and a p-value <0.05 was considered statistically significant. The statistical analyses were performed with R Studio 4.3.2.

## Results

### Study population

From four sites participating in the REFINE SPECT registry, we included 11035 participants who underwent SPECT/CT cardiac scans. After excluding cases that had incomplete T5-T11 scan coverage, missing clinical data, or had been used for EAT-model training, the final study group consisted of 9918 patients (Figure 2). The average computational time was 81.98±4.54 seconds per case.

**Figure 2.**
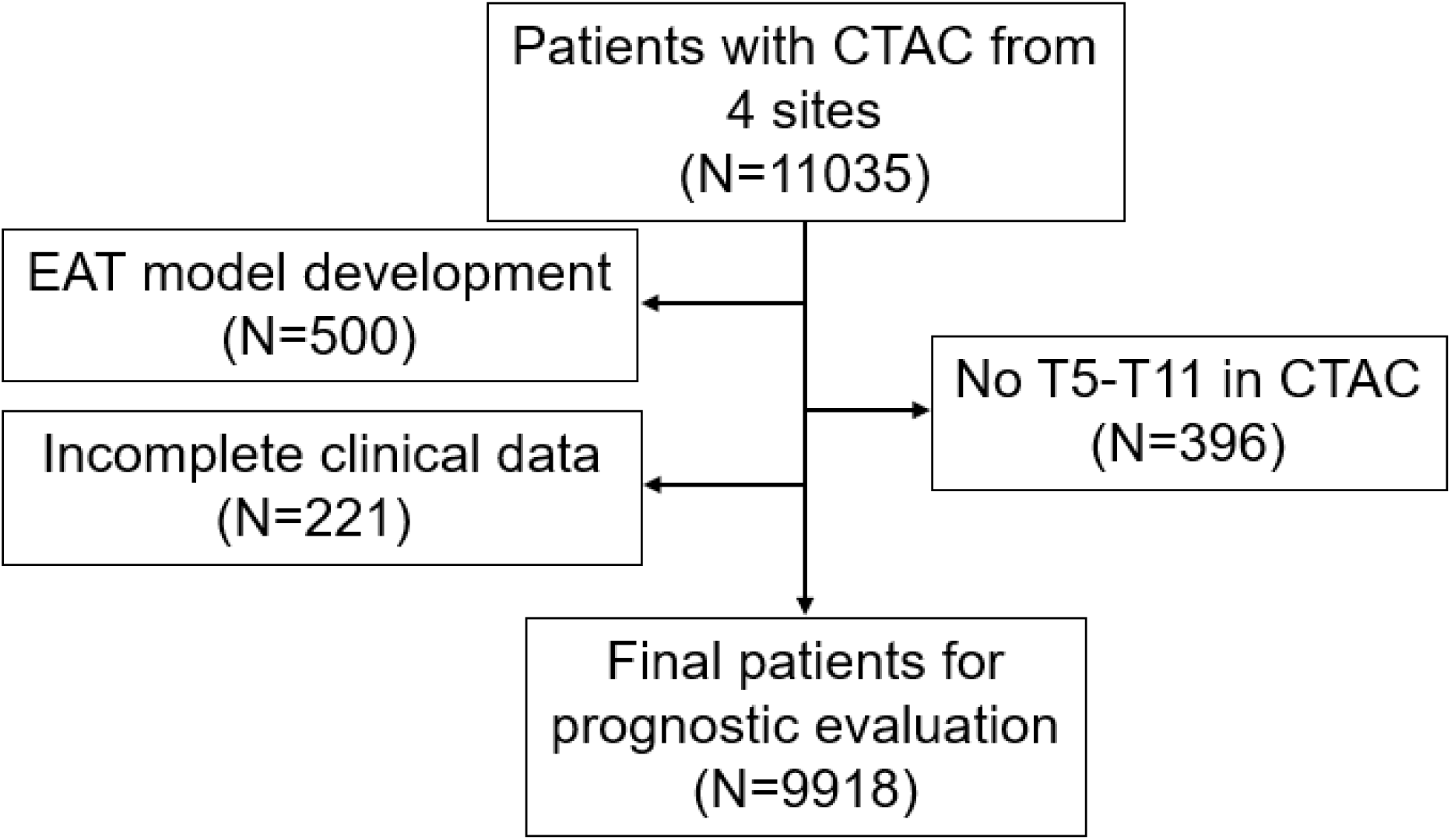
Study flowchart. Abbreviations: CTAC - computed tomography attenuation correction, EAT – epicardial adipose tissue, T – thoracic.

Of all included participants, 5451 (55%) were male and median age was 65 (57-73) years. Median follow-up time was 2.48 [1.45-3.60] years, during which 610 (6.2%) patients died (Table 1). Male patients had significantly higher bone (349 vs. 277, p<0.0001) and SM (914 vs. 674, p<0.0001) volume indices, but lower VAT (-86 vs. -82, p<0.0001) and EAT (-61 vs. -60, p=0.0097) attenuation as compared to females. Supplementary Tables 1-4 show baseline characteristics for all participants stratified by death, race (Black and White), age (<65 and ≥65 years), and BMI (<30 and ≥30 kg/m^2^), respectively. Youden index cutoffs for each measure are shown in Supplementary Table 5.

**Table 1.** Baseline characteristics for all participants stratified by sex.

|  | Male<br>(N=5451) | Female<br>(N=4467) | p-value |
| --- | --- | --- | --- |
| Age [years] | 64 (56, 73) | 66 (57, 74) | <0.0001 |
| BMI [kg/m<sup>2</sup>] | 30 (26, 34) | 31 (26, 36) | <0.0001 |
| Race |  |  | <0.0001 |
| American Indian or Alaska Native | 12 (0.2%) | 8 (0.2%) |  |
| Asian | 110 (2.0%) | 57 (1.3%) |  |
| Black or African American | 546 (10%) | 639 (14%) |  |
| Native Hawaiian or Other Pacific Islander | 6 (0.1%) | 13 (0.3%) |  |
| White | 2067 (38%) | 1296 (29%) |  |
| Unavailable | 2710 (50%) | 2454 (55%) |  |
| Hypertension | 3411 (63%) | 2756 (62%) | 0.3692 |
| Diabetes mellitus | 1623 (30%) | 1199 (27%) | 0.0013 |
| Dyslipidemia | 2996 (55%) | 2062 (46%) | <0.0001 |
| Smoking | 947 (17%) | 541 (12%) | <0.0001 |
| Family history of coronary artery disease | 1264 (23%) | 1198 (27%) | <0.0001 |
| Follow-up [years] | 2.40 (1.40, 3.66) | 2.57 (1.55, 3.64) | 0.0004 |
| Mortality | 382 (7.0%) | 228 (5.1%) | <0.0001 |
| Stress TPD [%] | 3 (1, 9) | 4 (1, 7) | 0.0050 |
| LVEF [%] | 59 (51, 66) | 70 (63, 78) | <0.0001 |
| Log(CAC score + 1) [AU] | 2.29 (0.00, 3.09) | 0.95 (0.00, 2.33) | <0.0001 |
| Bone attenuation [HU] | 258 (230, 287) | 253 (224, 289) | <0.0001 |
| Bone SD [HU] | 197 (178, 216) | 197 (176, 220) | 0.6175 |
| Bone volume index [cm<sup>3</sup>/m<sup>2</sup>] | 349 (312, 386) | 277 (249, 306) | <0.0001 |
| EAT attenuation [HU] | -61 (-68, -54) | -60 (-68, -53) | 0.0097 |
| EAT SD [HU] | 48 (44, 54) | 47 (42, 53) | <0.0001 |
| EAT volume index [cm<sup>3</sup>/m<sup>2</sup>] | 49 (35, 67) | 48 (34, 65) | 0.0295 |
| IMAT attenuation [HU] | -71 (-75, -67) | -69 (-73, -66) | <0.0001 |
| IMAT SD [HU] | 49 (38, 62) | 43 (35, 55) | <0.0001 |
| IMAT volume index [cm<sup>3</sup>/m<sup>2</sup>] | 101 (67, 155) | 74 (50, 110) | <0.0001 |
| VAT attenuation [HU] | -86 (-90, -81) | -82 (-86, -78) | <0.0001 |
| VAT SD [HU] | 57 (52, 65) | 57 (53, 64) | **0.6180** |
| VAT volume index [cm<sup>3</sup>/m<sup>2</sup>] | 451 (307, 619) | 310 (211, 432) | **<0.0001** |
| SAT attenuation [HU] | -99 (-102, -95) | -103 (-106, -99) | **<0.0001** |
| SAT SD [HU] | 41 (32, 51) | 37 (30, 48) | **<0.0001** |
| SAT volume index [cm<sup>3</sup>/m<sup>2</sup>] | 863 (583, 1243) | 1706 (1161, 2301) | **<0.0001** |
| SM attenuation [HU] | 33 (28, 38) | 26 (22, 31) | **<0.0001** |
| SM SD [HU] | 53 (42, 65) | 48 (40, 58) | **<0.0001** |
| SM volume index [cm<sup>3</sup>/m<sup>2</sup>] | 914 (768, 1060) | 674 (567, 789) | **<0.0001** |
| Ratio SM/ATs volume | 0.60 (0.45, 0.83) | 0.31 (0.24, 0.42) | **<0.0001** |

Values are presented as N (%) or median (IQ1, IQ3)
ATs – adipose tissue, BMI – body mass index, CAC – coronary artery calcium, CT – computed tomography, EAT – epicardial adipose tissue, HU – Hounsfield unit, IMAT – intramuscular adipose tissue, LVEF – left ventricular ejection fraction, MPI – myocardial perfusion imaging, SAT – subcutaneous adipose tissue, SD – standard deviation, SM – skeletal muscle, TPD – total perfusion deficit, VAT – visceral adipose tissue

### Body composition quantification predictors for all-cause mortality In entire study group

The quantitative predictors of mortality for six tissues are shown in Figure 3. The unadjusted and adjusted HR are presented in Table 2 and Supplementary Table 6 (extended information). High VAT attenuation (>-80 HU) was associated with an increased ACM risk (adjusted HR 2.39 [1.92, 2.96], p<0.0001). Patients with high bone attenuation were at lower risk of death (adjusted HR 0.77 [0.62, 0.95], p=0.0159). Likewise, high SM volume index was associated with a lower risk of death (adjusted HR 0.56 [0.44, 0.71], p<0.0001). In Supplementary Figures 3-5, we show risk stratification for six body composition attenuation measures stratified by cardiac variables (total perfusion deficit, left ventricular ejection fraction, and coronary artery calcium score). Figures 4 and 5 show different examples of body composition segmentation.

**Figure 3.**
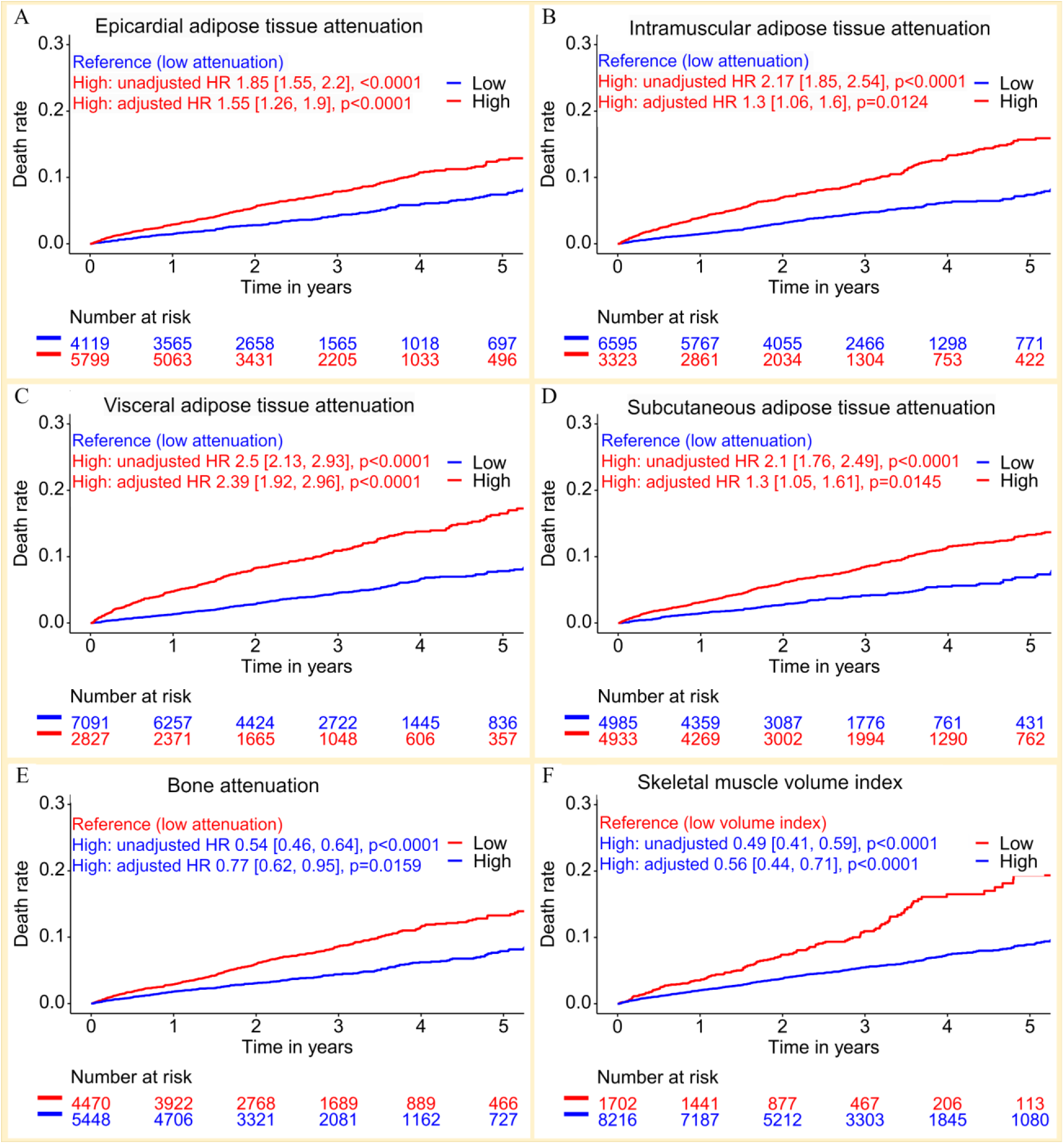
Kaplan-Meier curves stratified by body composition measures. A: epicardial adipose tissue (high attenuation: > -63 HU), B: intramuscular adipose tissue (high attenuation: > -68 HU), C: visceral adipose tissue (high attenuation: > -80 HU), D: subcutaneous adipose tissue (high attenuation: > -101 HU), E: bone (high attenuation: > 250 HU), F: skeletal muscle (high volume index: > 597.16 cm^3^/m^2^). Hazard ratios (HR) are shown (both unadjusted and adjusted for 11 clinical and imaging variables and other 18 body composition measures).

**Figure 4.**
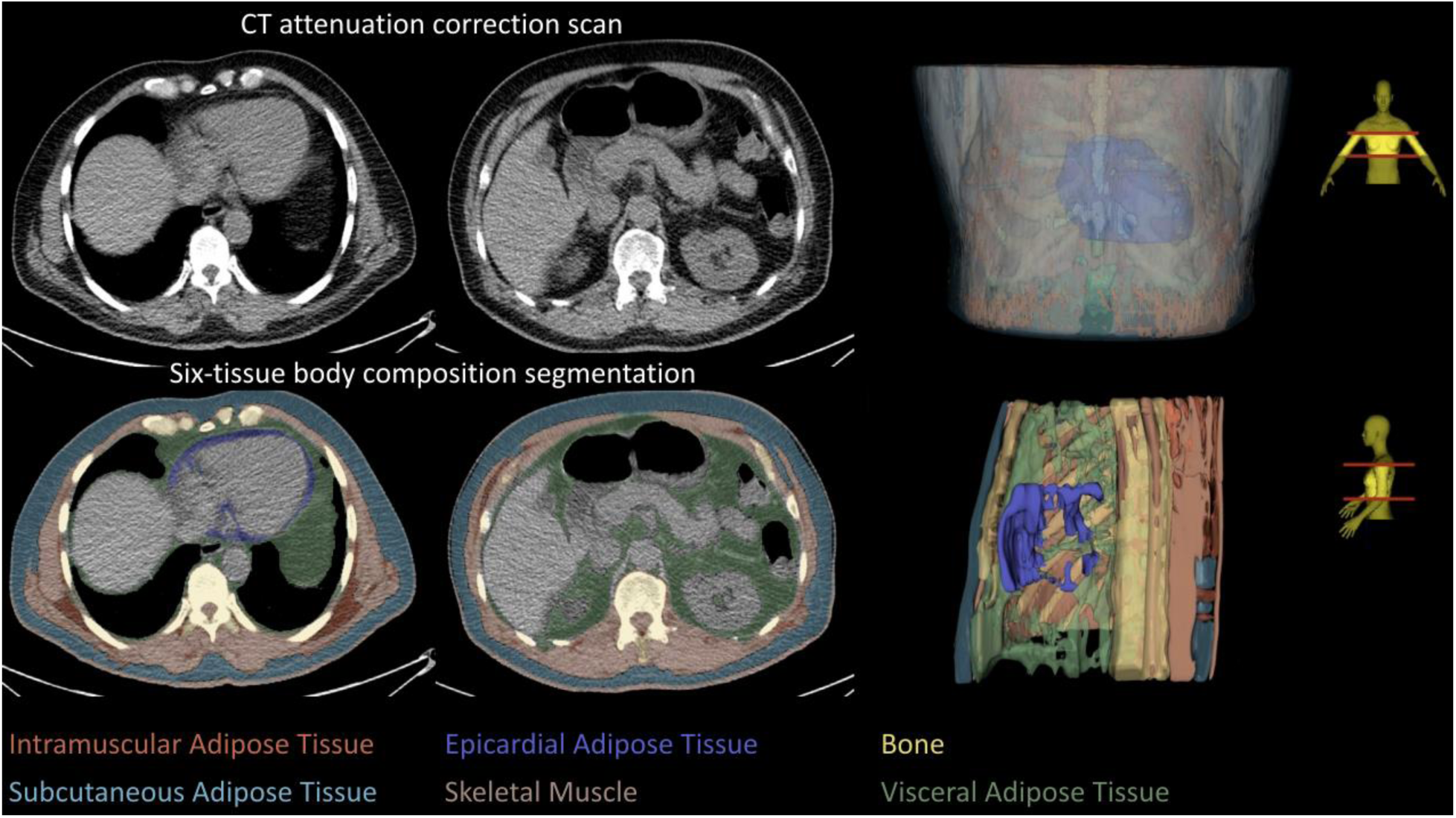
Example of body composition segmentation from computed tomography attenuation correction scans in a male patient with a body mass index of 26.4 kg/m^2^.

**Figure 5.**
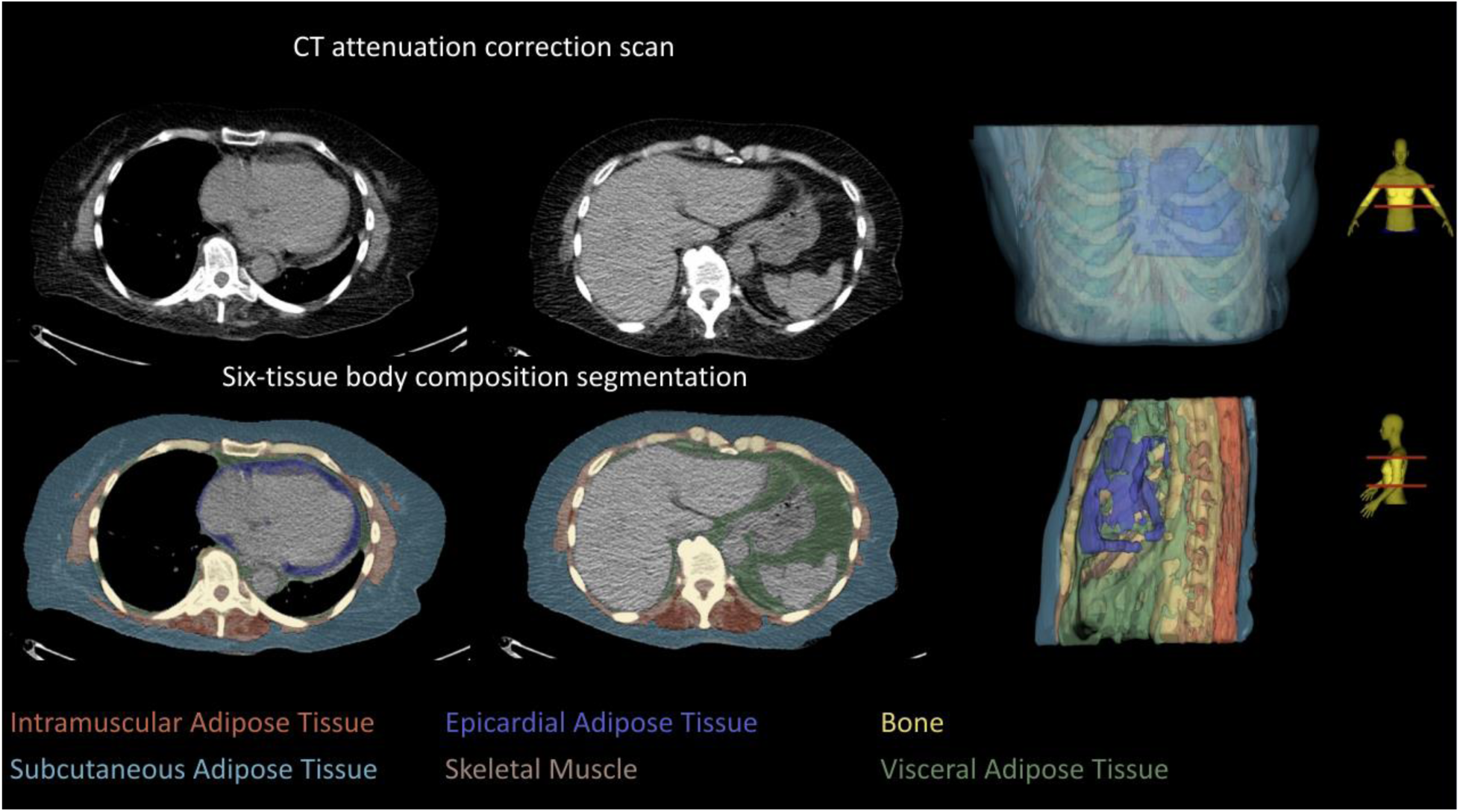
Example of body composition segmentation from computed tomography attenuation correction scans in a female patient with a body mass index of 25.8 kg/m^2^.

**Table 2.** Comparison between unadjusted and adjusted hazard ratios. Lower values are considered reference.

| Body Composition Quantification | Hazard Ratios |  |  |  |
| --- | --- | --- | --- | --- |
|  | Univariable |  | Multivariable* |  |
|  | HR [95% CI] | P-value | HR [95% CI] | P-value |
| Bone attenuation [HU] | **0.54 [0.46, 0.64]** | **<0.0001** | **0.77 [0.62, 0.95]** | **0.0159** |
| Bone SD [HU] | **0.79 [0.67, 0.93]** | **0.0039** | 1.10 [0.89, 1.36] | 0.3911 |
| Bone volume index [cm<sup>3</sup>/m<sup>2</sup>] | **1.34 [1.14, 1.59]** | **0.0005** | 1.16 [0.93, 1.46] | 0.1922 |
| EAT attenuation [HU] | **1.85 [1.55, 2.20]** | **<0.0001** | **1.55 [1.26, 1.90]** | **<0.0001** |
| EAT SD [HU] | **1.62 [1.37, 1.90]** | **<0.0001** | **1.50 [1.23, 1.83]** | **<0.0001** |
| EAT volume index [cm<sup>3</sup>/m<sup>2</sup>] | **1.70 [1.44, 1.99]** | **<0.0001** | **1.56 [1.28, 1.91]** | **<0.0001** |
| IMAT attenuation [HU] | **2.17 [1.85, 2.54]** | **<0.0001** | **1.30 [1.06, 1.60]** | **0.0124** |
| IMAT SD [HU] | **0.71 [0.54, 0.93]** | **0.0123** | 0.90 [0.65, 1.26] | 0.5485 |
| IMAT volume index [cm<sup>3</sup>/m<sup>2</sup>] | **1.33 [1.12, 1.57]** | **<0.0009** | **1.29 [1.03, 1.62]** | **0.0165** |
| VAT attenuation [HU] | **2.50 [2.13, 2.93]** | **<0.0001** | **2.39 [1.92, 2.96]** | **<0.001** |
| VAT SD [HU] | 0.89 [0.76, 1.05] | 0.1642 | **0.78 [0.63, 0.96]** | **0.0219** |
| VAT volume index [cm<sup>3</sup>/m<sup>2</sup>] | 1.12 [0.94, 1.32] | 0.2072 | 1.04 [0.83, 1.31] | 0.7274 |
| SAT attenuation [HU] | **2.10 [1.76, 2.49]** | **<0.0001** | **1.30 [1.05, 1.61]** | **0.0145** |
| SAT SD [HU] | 1.07 [0.90, 1.28] | 0.4458 | 1.01 [0.79, 1.29] | 0.9502 |
| SAT volume index [cm<sup>3</sup>/m<sup>2</sup>] | **0.60 [0.51, 0.70]** | **<0.0001** | 0.94 [0.74, 1.20] | 0.6429 |
| SM attenuation [HU] | **0.61 [0.51, 0.72]** | **<0.0001** | **0.54 [0.44, 0.67]** | **<0.0001** |
| SM SD [HU] | **1.67 [1.41, 1.99]** | **<0.0001** | **1.64 [1.27, 2.12]** | **0.0002** |
| SM volume index [cm<sup>3</sup>/m<sup>2</sup>] | **0.49 [0.41, 0.59]** | **<0.0001** | **0.56 [0.44, 0.71]** | **<0.0001** |
| Ratio SM/ATs volume | **1.24 [1.04, 1.49]** | **0.0153** | 1.15 [0.90, 1.48] | 0.2671 |

Bold indicates statistical significance.
\*Adjusted for clinical, perfusion, and other body composition factors: sex, age, body mass index, hypertension, diabetes mellitus, dyslipidemia, family history of coronary artery disease, smoking, stress total perfusion deficit, left ventricle ejection fraction, and deep-learning-coronary artery calcium score and all the other body composition quantifications.
CI – confidence interval, EAT – epicardial adipose tissue, IMAT – intramuscular adipose tissue, HU – Hounsfield units, SAT – subcutaneous adipose tissue, SD – standard deviation, VAT – visceral adipose tissue, ratio SM/ATs volume – ratio of skeletal muscle volume to all adipose tissues volume.

### In subgroups

VAT attenuation was a strong predictor for ACM in all subgroups distinguished based on sex, race, age, and BMI categories. High VAT attenuation (>-80 HU) was associated with an elevated ACM risk in both male and female participants (unadjusted HRs 2.79 [2.28, 3.42], p<0.0001 and 2.71 [2.08, 3.54], p<0.0001 [Supplementary Table 7]; HRs adjusted for 10 clinical and imaging factors 2.42 [1.94, 3.04], p<0.0001 and 2.28 [1.70, 3.07], p<0.0001 [Supplementary Table 8]; HRs additionally adjusted for remaining 18 body composition measures 2.43 [1.82, 3.24], p<0.0001 and 2.40 [1.68, 3.43], p<0.0001 [Supplementary Table 9]).

High SAT attenuation significantly increased death risk in both female and male patients (unadjusted HR 3.22 [2.46, 4.21], p<0.0001; 1.37 [1.09, 1.72], p=0.0069, respectively) (Supplementary Table 7). High VAT attenuation elevated the risk of mortality in all gender groups (male and female) (unadjusted HRs 2.79 [2.28, 3.42], p<0.0001; 2.71 [2.08, 3.54], p<0.0001, respectively) (Supplementary Table 7), White and Black groups (unadjusted HRs 2.91 [2.19, 3.87], p<0.0001; 1.99 [1.26, 3.15], p=0.0032, respectively) (Supplementary Table 10), all age groups (<65 years and ≥65 years) (unadjusted HRs 2.97 ([2.22, 3.97], p<0.0001) and 2.22 ([1.85, 2.67], p<0.0001), respectively) (Supplementary Table 11), and all BMI groups (<30 kg/m^2^ and ≥30 kg/m^2^) (unadjusted HRs 2.33 [1.89, 2.88], p<0.0001; 2.24 [1.70-2.97], p<0.0001, respectively) (Supplementary Table 12). For subjects with BMI ≥30 kg/m^2^, high EAT, IMAT, and VAT volume indexes were at elevated risk of death as compared to patients with low volume indices (unadjusted HRs 2.36 [1.78-3.13], p<0.0001; 2.38 [1.58-3.58], p<0.0001; 1.56 [1.21-2.01], p=0.0006, respectively) (Supplementary Table 12).

## Discussion

We developed AI-based volumetric CTAC-body composition segmentation in patients undergoing cardiac perfusion imaging. This simple and automatic approach reveals robust information on body tissue composition available in all CTAC scans, but not utilized to date. This task was accomplished by integrating fully automated segmentation of skeletal muscles, bone, subcutaneous, intramuscular, and visceral adipose tissues with our previously validated deep-learning-model for EAT quantification from CTAC maps in a fast (less than average 2 minutes per case) and reproducible manner with no additional exposure to radiation. The algorithm was applied to scans with different image acquisition parameters acquired with various scanners from different manufacturers demonstrating wide generalizability. We showed strong and independent prognostic value for several body composition measures, which remained predictive of all-cause mortality even after adjustment for key clinical variables and imaging variables. These body composition measures are prognostically complementary to each other and to major cardiac perfusion measures.

Patients with abnormal body composition measurements have worse outcomes and increased mortality,^3, 14^ which was also demonstrated in our study. Abnormal body composition phenotypes include low muscle mass (sarcopenia),^15^ high fat mass (obesity),^4^ and both low muscle and fat mass (cachexia).^16^ Obesity is an alarmingly increasing global public health problem^17^ that affects the psychosocial and physical aspects of quality of life and is associated with various comorbidities such as cardiovascular disease^18^ and mortality,^19^ but BMI-based assessment does not provide adequate information.^2^ Sarcopenia and cachexia are also recognized to be associated with negative outcomes like physical disability,^20^ prolonged hospitalizations,^21^ or death.^15, 22^ Utilizing the tools developed in our study, all these prevalent conditions relevant to cardiometabolic health can be monitored comprehensively and automatically in any patients undergoing scanning which includes low-dose chest CT imaging.

Body tissue composition analysis provides information of vital clinical importance. Prior studies utilized mainly abdominal CT scans and a single CT slice for segmentation^6, 7^ of a limited number of body tissues (skeletal muscle, visceral or subcutaneous adipose tissue).^2, 5–7^ Xu et al. demonstrated the predictive value of body composition measurements from low-dose chest CT scans for mortality prediction in smokers from the National Lung Screening Trial (NLST), although analyzing only a limited number (n=3) of specific slices.^5^ Our group studied the association of volumetric EAT quantification from CTAC with adverse events.^8^ To our knowledge, this is the first comprehensive volumetric body composition analysis of six tissues simultaneously from chest CTAC maps (which are of even lower quality than standard lung CT scans). These maps are utilized for attenuation correction and visual estimation of calcium during hybrid imaging, yet they contain other valuable information that is not currently considered during clinical reporting.

Importantly, to provide a comprehensive assessment, we also included EAT analysis, which is known to be associated with the risk of cardiovascular events.^8, 23^ In our holistic approach, we evaluated measures of automatically segmented tissues inside/outside the thoracic cavity within automatically-determined sub-volume defined by T5-T11 vertebrae. Among all the volumetric measures, tissue density (defined by attenuation in HU) remained the strongest mortality predictor when adjusted for other variables, for bone, VAT, SAT, and SM. VAT attenuation was a strong mortality predictor across all subgroups. Since excess adipose tissue, particularly in the visceral compartment (visceral obesity), is associated with metabolic syndrome, cardiovascular disease, and increased mortality,^24, 25^ more comprehensive cross-sectional image analysis may be important. This analysis can be easily performed with AI-assistance from previously obtained scans, without the need for extra radiation or imaging time.

Our study has several limitations. We performed only a regional and volumetric body composition analysis (T5-T11) to predict all-cause mortality, which was done to ensure standard coverage for the vast majority of patients undergoing the MPI scans. Our segmentation framework was a multi-stage approach that required the segmentation results from deep-learning-based segmentation models, potentially increasing the running time; however, the total end-to-end time of 2 minutes should be clinically feasible as the analysis can be performed in background. We did not distinguish the intermuscular and intramuscular adipose tissue while analyzing IMAT. Similarly, compact bone and bone marrow were not analyzed separately. We did not manually annotate segmentations for evaluating segmentation algorithm due to the large scale of our cohort. Lastly, our study only considered quantification of different tissues from chest CT scan, but body composition quantification from abdominal CT could potentially provide additive prognostic value.^2^

In conclusion, we demonstrated the utility of performing opportunistic volumetric body composition analysis for prognostic prediction of all-cause mortality from CTAC in patients undergoing cardiac perfusion imaging by developing a simple and annotation-free body composition segmentation framework for six tissues. The obtained volumetric body composition measurements offer additional prognostic value over perfusion imaging, calcium measures and clinical data and can be performed automatically using data from existing scans without additional radiation or imaging time.

## Contributors

JY developed the algorithms, conducted data processing/experiments/analysis, and co-wrote the manuscript. AM co-wrote the manuscript and contributed materials, clinical expertise, and technical expertise. PJS designed the study, provided the overall guidance and study funding, co-wrote the manuscript, and contributed materials, clinical expertise, and technical expertise. AS, RJM, JG, WZ, AK, NM, ML, JZ, VB, JXL, TDR, AJE, AF, EJM, AJS, JK, PK, DD, and DSB contributed materials, clinical expertise, and technical expertise. All authors critically revised the manuscript and contributed to its formation. JY, AM, AS, and JG directly accessed and verified the data in the study. All authors had full access to all the data in the study, accept the final responsibility to submit for publication and take responsibility for the contents of the manuscript.

## Data sharing

All implementation for the body composition approach used primarily Python-based image processing libraries including Python version 3.11.5, Scikit-image version 0.20.0 (https://scikit-image.org/), OpenCV version 4.6.0 (https://opencv.org/), and SciPy version 1.11.3 (https://scipy.org/). TotalSegmentator v2 (https://github.com/wasserth/TotalSegmentator), EAT segmentation model, and CAC segmentation model were implemented in PyTorch.^8,11^ All the statistical analysis was performed using RStudio 4.3.2, and the R libraries used include dplyr, tidyr, readxl, stringr, corrplot, survival, survminer, VennDiagram, gtsummary, gt, forestmodel, tidyverse, tidytidbits, adjustedCurves, and survivalAnalysis. All the experiments were completed using a desktop with Windows 10 Pro 64-bit operating system, 256GB RAM, AMD Ryzen Threadripper 3950X 24-Core Processor CPU, and NVIDIA TITAN RTX 24 GB GPU. The source code is available upon reasonable written request. Access to the code is available from the date of publication.

## Data Availability

All implementation for the body composition approach used primarily Python-based image processing libraries including Python version 3.11.5, Scikit-image version 0.20.0 (https://scikit-image.org/), OpenCV version 4.6.0 (https://opencv.org/), and SciPy version 1.11.3 (https://scipy.org/). TotalSegmentator v2 (https://github.com/wasserth/TotalSegmentator), EAT segmentation model, and CAC segmentation model were implemented in PyTorch.8,11 All the statistical analysis was performed using RStudio 4.3.2, and the R libraries used include dplyr, tidyr, readxl, stringr, corrplot, survival, survminer, VennDiagram, gtsummary, gt, forestmodel, tidyverse, tidytidbits, adjustedCurves, and survivalAnalysis. All the experiments were completed using a desktop with Windows 10 Pro 64-bit operating system, 256GB RAM, AMD Ryzen Threadripper 3950X 24-Core Processor CPU, and NVIDIA TITAN RTX 24 GB GPU. The source code is available upon reasonable written request. Access to the code is available from the date of publication.

## Acknowledgements

This research was supported in part by Grants R01HL089765 and R35HL161195 from the National Heart, Lung, and Blood Institute/National Institutes of Health (NHLBI/NIH) (PI: Piotr Slomka).

## List of abbreviations

ACM: all-cause mortality
AI: artificial intelligence
BMI: body mass index
CAC: coronary artery calcium
CTAC: computed tomography attenuation correction
EAT: epicardial adipose tissue
IMAT: intramuscular adipose tissue
HU: Hounsfield unit
LVEF: left ventricular ejection fraction
MPI: myocardial perfusion imaging
SAT: subcutaneous adipose tissue
SM: skeletal muscle
TPD: total perfusion deficit
VAT: visceral adipose tissue

