## Supplemental Material for "AI-based volumetric six-tissue body composition quantification from CT cardiac attenuation scans enhances mortality prediction: multicenter study"

**Supplementary appendix**

**Supplementary methods**

The proposed annotation-free and image processing-based body composition segmentation framework includes our body composition segmentation approach, an existing publicly available trained TotalSegmentator v2 model, an existing trained EAT segmentation model, and an existing image processing-based body segmentation module.^1-3^ We first separate the body parts inside the rib cage from those outside the rib cage by exploiting segmentations from TotalSegmentator v2 (<https://github.com/wasserth/TotalSegmentator>) and image processing techniques. We then apply thresholding to differentiate between the bone, the muscle, and the adipose tissue which had different Hounsfield Unit (HU) ranges.

Our proposed body composition segmentation approach consists of two core modules: the image rib cage segmentation module, and the body composition module for segmenting 5 tissues (bone, SM, SAT, IMAT, and VAT). The rib cage segmentation module was based on the principle that the rib cage region was convex in the sagittal view and the coronal view, and locally-convex in the axial view. The TotalSegmentator v2 provided segmentations of 117 different structures of human body including organs inside the rib cage such as lung and spleen. The pre-segmented organs allowed us to use convex hull of the organs inside the rib cage region to estimate the rib cage region.

Utilizing rib-cage segmentation, we could define the VAT via thresholding inside rib-cage volume. Outside the rib cage, we separated the bone, the SM, and the adipose tissue by thresholding due to the distinct differences in their HU ([151, 1500] for bone, [-29, 150] for skeletal muscle (SM), and [-190,-30] for adipose tissue). For the adipose tissue outside the rib cage, we utilized location information (whether the adipose tissue component was touching the skin or was surrounded by or infiltrating muscle) to separate the subcutaneous adipose tissue (SAT) from the intramuscular adipose tissue (IMAT). By combining the EAT mask from the existing trained EAT segmentation model, we could further separate the VAT into EAT and non-epicardial VAT.^2^ All of our implementations used basic image processing techniques supported by generic Python libraries such as Scikit-image, OpenCV, and SciPy. More details of our body composition segmentation approach can be found in Supplementary Figure 2.

**Supplementary Tables**

**Supplementary Table 1. Baseline characteristics for all participants stratified by all-cause mortality**

|  | **Death**  **N=610** | **Non-Death**  **N=9308** | **p-value** |
| --- | --- | --- | --- |
| Male | 382 (63%) | 5,069 (54%) | <0.001 |
| Age [years] | 71 (63, 78) | 65 (56, 73) | <0.001 |
| BMI [kg/m^2^] | 28 (24, 33) | 30 (26, 35) | <0.001 |
| Race |  |  | 0.024 |
| American Indian or Alaska Native | 3 (0.5%) | 17 (0.2%) |  |
| Asian | 4 (0.7%) | 163 (1.8%) |  |
| Black or African American | 77 (13%) | 1,108 (12%) |  |
| Native Hawaiian or Other Pacific Islander | 3 (0.5%) | 16 (0.2%) |  |
| White | 192 (31%) | 3,171 (34%) |  |
| Unavailable | 331 (54%) | 4,833 (52%) |  |
| Hypertension | 395 (65%) | 5,772 (62%) | 0.180 |
| Diabetes mellitus | 226 (37%) | 2,596 (28%) | <0.001 |
| Dyslipidemia | 314 (51%) | 4,744 (51%) | 0.810 |
| Smoking | 106 (17%) | 1,382 (15%) | 0.090 |
| Family history of coronary artery disease | 127 (21%) | 2,335 (25%) | 0.018 |
| Follow-up [years] | 1.56 (0.64, 2.82) | 2.55 (1.52, 3.70) | <0.001 |
| Stress TPD [%] | 5 (2, 11) | 3 (1, 8) | <0.001 |
| LVEF [%] | 58 (44, 68) | 64 (56, 72) | <0.001 |
| Log(CAC score+1) [AU] | 2.59 (1.52, 3.25) | 1.67 (0.00, 2.81) | <0.001 |
| Bone attenuation [HU] | 241 (212, 276) | 257 (228, 288) | <0.001 |
| Bone SD [HU] | 191 (171, 211) | 197 (177, 218) | <0.001 |
| Bone volume index [cm^3^/m^2^] | 329 (284, 382) | 311 (273, 358) | <0.001 |
| EAT attenuation [HU] | -58 (-66, -53) | -61 (-68, -54) | <0.001 |
| EAT SD [HU] | 49 (44, 55) | 48 (43, 53) | <0.001 |
| EAT volume index [cm^3^/m^2^] | 55 (37, 75) | 48 (34, 66) | <0.001 |
| IMAT attenuation [HU] | -67 (-73, -64) | -70 (-74, -67) | <0.001 |
| IMAT SD [HU] | 46 (36, 59) | 46 (37, 60) | 0.7 |
| IMAT volume index [cm^3^/m^2^] | 91 (60, 152) | 87 (57, 134) | 0.01 |
| VAT attenuation [HU] | -80 (-86, -74) | -85 (-89, -80) | <0.001 |
| VAT SD [HU] | 56 (52, 64) | 57 (52, 65) | 0.008 |
| VAT volume index [cm^3^/m^2^] | 381 (254, 550) | 380 (253, 533) | 0.47 |
| SAT attenuation [HU] | -98 (-102, -92) | -101 (-104, -97) | <0.001 |
| SAT SD [HU] | 39 (32, 49) | 39 (31, 50) | 0.66 |
| SAT volume index [cm^3^/m^2^] | 977 (607, 1,533) | 1,171 (728, 1,804) | <0.001 |
| SM attenuation [HU] | 28 (23, 33) | 30 (25, 35) | <0.001 |
| SM SD [HU] | 52 (42, 62) | 50 (41, 62) | 0.024 |
| SM volume index [cm^3^/m^2^] | 764 (595, 920) | 796 (651, 972) | <0.001 |
| Ratio SM/ATs volume | 0.46 (0.32, 0.71) | 0.45 (0.31, 0.68) | 0.085 |

Values are presented as N (%) or median (IQ1, IQ3)

AU – Agatston unit, ATs – adipose tissues, BMI – body mass index, CAC – coronary artery calcium, CAD – coronary artery disease, CT – computed tomography, EAT – epicardial adipose tissue, HU – Hounsfield unit, IMAT – intramuscular adipose tissue, LVEF – left ventricular ejection fraction, MPI – myocardial perfusion imaging, Ratio SM/ATs volume – ratio of skeletal muscle volume to all adipose tissues volume, SAT – subcutaneous adipose tissue, SD – standard deviation, SM – skeletal muscle, TPD – total perfusion deficit, VAT – visceral adipose tissue

**Supplementary Table 2. Baseline characteristics for all participants stratified by Black or White in the population where race information was available.**

|  | **Black or African American**  **(N=1185)** | **White**  **(N=3363)** | **p-value** |
| --- | --- | --- | --- |
| Age [years] | 60 (52, 67) | 66 (58, 74) | <0.001 |
| BMI [kg/m^2^] | 32 (27, 37) | 30 (26, 34) | <0.001 |
| Male | 546 (46%) | 2,067 (61%) | <0.001 |
| Hypertension | 825 (70%) | 2,001 (60%) | <0.001 |
| Diabetes mellitus | 440 (37%) | 782 (23%) | <0.001 |
| Dyslipidemia | 525 (44%) | 1,763 (52%) | <0.001 |
| Smoking | 264 (22%) | 616 (18%) | 0.003 |
| Family history of CAD | 105 (8.9%) | 463 (14%) | <0.001 |
| Follow-up [years] | 2.49 (1.36, 3.72) | 2.30 (1.33, 3.42) | 0.003 |
| Mortality | 77 (6.5%) | 192 (5.7%) | 0.32 |
| Stress TPD [%] | 2.9 (1.0, 6.2) | 2.5 (0.9, 5.9) | 0.11 |
| LVEF [%] | 62 (53, 69) | 65 (56, 72) | <0.001 |
| Log(CAC score+1) [AU] | 0.99 (0.00, 2.34) | 2.25 (0.00, 3.07) | <0.001 |
| Bone attenuation [HU] | 296 (263, 331) | 255 (228, 283) | <0.001 |
| Bone SD [HU] | 214 (193, 235) | 204 (185, 223) | <0.001 |
| Bone volume index [cm^3^/m^2^] | 284 (249, 325) | 310 (272, 357) | <0.001 |
| EAT attenuation [HU] | -55 (-65, -48) | -62 (-69, -55) | <0.001 |
| EAT SD [HU] | 49 (44, 57) | 50 (44, 59) | <0.001 |
| EAT volume index [cm^3^/m^2^] | 35 (25, 48) | 54 (39, 72) | <0.001 |
| IMAT attenuation [HU] | -71 (-76, -66) | -70 (-74, -67) | 0.11 |
| IMAT SD [HU] | 52 (39, 68) | 53 (39, 67) | 0.42 |
| IMAT volume index [cm^3^/m^2^] | 93 (59, 152) | 90 (60, 142) | 0.35 |
| VAT attenuation [HU] | -81 (-85, -76) | -85 (-90, -80) | <0.001 |
| VAT SD [HU] | 65 (58, 71) | 63 (56, 69) | <0.001 |
| VAT volume index [cm^3^/m^2^] | 262 (179, 368) | 386 (265, 530) | <0.001 |
| SAT attenuation [HU] | -101 (-104, -96) | -102 (-105, -98) | <0.001 |
| SAT SD [HU] | 46 (33, 57) | 46 (34, 57) | 0.97 |
| SAT volume index [cm^3^/m^2^] | 1,354 (741, 2,095) | 1,042 (671, 1,592) | <0.001 |
| SM attenuation [HU] | 30 (25, 35) | 30 (25, 35) | 0.095 |
| SM SD [HU] | 55 (43, 69) | 57 (43, 69) | 0.035 |
| SM volume index [cm^3^/m^2^] | 826 (687, 1,014) | 760 (607, 928) | <0.001 |
| Ratio SM/ATs volume | 0.46 (0.32, 0.76) | 0.46 (0.31, 0.67) | 0.057 |

Values are presented as N (%) or median (IQ1, IQ3)

ATs – adipose tissues, BMI – body mass index, CAC – coronary artery calcium, CAD – coronary artery disease, CT – computed tomography, EAT – epicardial adipose tissue, HU – Hounsfield unit, IMAT – intramuscular adipose tissue, LVEF – left ventricular ejection fraction, MPI – myocardial perfusion imaging, SAT – subcutaneous adipose tissue, SD – standard deviation, SM – skeletal muscle, TPD – total perfusion deficit, VAT – visceral adipose tissue, Ratio SM/ATs volume – ratio of SM volume to AT volume

**Supplementary Table 3. Baseline characteristics for all participants stratified by age (<65 and ≥65 years)**

|  | **<65 years**  **(N=4819)** | **≥65 years**  **(N=5099)** | **p-value** |
| --- | --- | --- | --- |
| Male | 2,747 (57%) | 2,704 (53%) | <0.001 |
| BMI [kg/m^2^] | 31 (27, 36) | 29 (26, 34) | <0.001 |
| Race |  |  | <0.001 |
| American Indian or Alaska Native | 12 (0.2%) | 8 (0.2%) |  |
| Asian | 93 (1.9%) | 74 (1.5%) |  |
| Black or African American | 800 (17%) | 385 (7.6%) |  |
| Native Hawaiian or Other Pacific Islander | 11 (0.2%) | 8 (0.2%) |  |
| White | 1,478 (31%) | 1,885 (37%) |  |
| Unavailable | 2,425 (50%) | 2,739 (54%) |  |
| Hypertension | 2,658 (55%) | 3,509 (69%) | <0.001 |
| Diabetes mellitus | 1,293 (27%) | 1,529 (30%) | <0.001 |
| Dyslipidemia | 2,130 (44%) | 2,928 (57%) | <0.001 |
| Smoking | 879 (18%) | 609 (12%) | <0.001 |
| Family history of coronary artery disease | 1,324 (27%) | 1,138 (22%) | <0.001 |
| Follow-up [years] | 2.49 (1.43, 3.70) | 2.48 (1.50, 3.62) | 0.69 |
| Mortality | 174 (3.6%) | 436 (8.6%) | <0.001 |
| Stress TPD [%] | 3 (1, 7) | 4 (1, 9) | <0.001 |
| LVEF [%] | 63 (56, 71) | 65 (55, 73) | <0.001 |
| Log(CAC score+1) [AU] | 0.00 (0.00, 2.24) | 2.41 (1.14, 3.09) | <0.001 |
| Bone attenuation [HU] | 268 (240, 299) | 244 (217, 274) | <0.001 |
| Bone SD [HU] | 200 (181, 221) | 193 (173, 214) | <0.001 |
| Bone volume index [cm^3^/m^2^] | 306 (269, 349) | 320 (279, 369) | <0.001 |
| EAT attenuation [HU] | -59 (-67, -52) | -62 (-69, -55) | <0.001 |
| EAT SD [HU] | 48 (43, 54) | 48 (43, 54) | 0.7 |
| EAT volume index [cm^3^/m^2^] | 43 (30, 61) | 54 (39, 72) | <0.001 |
| IMAT attenuation [HU] | -72 (-76, -68) | -69 (-73, -65) | <0.001 |
| IMAT SD [HU] | 48 (38, 62) | 44 (36, 57) | <0.001 |
| IMAT volume index [cm^3^/m^2^] | 88 (57, 141) | 87 (58, 129) | 0.15 |
| VAT attenuation [HU] | -85 (-89, -79) | -84 (-88, -79) | <0.001 |
| VAT SD [HU] | 59 (54, 67) | 55 (52, 63) | <0.001 |
| VAT volume index [cm^3^/m^2^] | 349 (229, 495) | 410 (277, 577) | <0.001 |
| SAT attenuation [HU] | -101 (-104, -97) | -100 (-104, -96) | <0.001 |
| SAT SD [HU] | 41 (32, 52) | 38 (30, 48) | <0.001 |
| SAT volume index [cm^3^/m^2^] | 1,182 (715, 1,856) | 1,137 (722, 1,741) | 0.017 |
| SM attenuation [HU] | 32 (27, 37) | 28 (24, 33) | <0.001 |
| SM SD [HU] | 52 (41, 64) | 50 (41, 60) | <0.001 |
| SM volume index [cm^3^/m^2^] | 853 (705, 1,036) | 741 (607, 897) | <0.001 |
| Ratio SM/ATs volume | 0.50 (0.33, 0.76) | 0.42 (0.30, 0.61) | <0.001 |

Values are presented as N (%) or median (IQ1, IQ3)

AU – Agatston unit, ATs – adipose tissues, BMI – body mass index, CAC – coronary artery calcium, CAD – coronary artery disease, CT – computed tomography, EAT – epicardial adipose tissue, HU – Hounsfield unit, IMAT – intramuscular adipose tissue, LVEF – left ventricular ejection fraction, MPI – myocardial perfusion imaging, Ratio SM/ATs volume – ratio of skeletal muscle volume to all adipose tissues volume, SAT – subcutaneous adipose tissue, SD – standard deviation, SM – skeletal muscle, TPD – total perfusion deficit, VAT – visceral adipose tissue

**Supplementary Table 4. Baseline characteristics for all participants stratified by BMI (<30 and ≥30 kg/m^2^)**

|  | **BMI <30 kg/m^2^**  **(N=4845)** | **BMI ≥30 kg/m^2^**  **(N=5073)** | **p-value** |
| --- | --- | --- | --- |
| Age [years] | 67 (58, 75) | 63 (55, 71) | <0.001 |
| Male | 2,836 (59%) | 2,615 (52%) | <0.001 |
| Race |  |  | <0.001 |
| American Indian or Alaska Native | 11 (0.2%) | 9 (0.2%) |  |
| Asian | 119 (2.5%) | 48 (0.9%) |  |
| Black or African American | 481 (9.9%) | 704 (14%) |  |
| Native Hawaiian or Other Pacific Islander | 10 (0.2%) | 9 (0.2%) |  |
| White | 1,755 (36%) | 1,608 (32%) |  |
| Unavailable | 2,469 (51%) | 2,695 (53%) |  |
| Hypertension | 2,736 (56%) | 3,431 (68%) | <0.001 |
| Diabetes mellitus | 1,128 (23%) | 1,694 (33%) | <0.001 |
| Dyslipidemia | 2,381 (49%) | 2,677 (53%) | <0.001 |
| Smoking | 735 (15%) | 753 (15%) | 0.65 |
| Family history of coronary artery disease | 1,220 (25%) | 1,242 (24%) | 0.42 |
| Follow-up [years] | 2.38 (1.40, 3.64) | 2.56 (1.54, 3.68) | <0.001 |
| Mortality | 367 (7.6%) | 243 (4.8%) | <0.001 |
| Stress TPD [%] | 3 (1, 7) | 4 (1, 9) | <0.001 |
| LVEF [%] | 64 (55, 72) | 64 (56, 72) | 0.52 |
| Log(CAC score+1) [AU] | 1.92 (0.00, 2.95) | 1.56 (0.00, 2.72) | <0.001 |
| Bone attenuation [HU] | 256 (228, 287) | 255 (226, 288) | 0.43 |
| Bone SD [HU] | 197 (178, 219) | 196 (176, 217) | 0.007 |
| Bone volume index [cm^3^/m^2^] | 304 (266, 349) | 321 (281, 368) | <0.001 |
| EAT attenuation [HU] | -59 (-67, -53) | -62 (-69, -54) | <0.001 |
| EAT SD [HU] | 46 (42, 51) | 50 (44, 55) | <0.001 |
| EAT volume index [cm^3^/m^2^] | 42 (30, 57) | 56 (40, 76) | <0.001 |
| IMAT attenuation [HU] | -67 (-71, -64) | -73 (-77, -69) | <0.001 |
| IMAT SD [HU] | 42 (33, 53) | 51 (39, 65) | <0.001 |
| IMAT volume index [cm^3^/m^2^] | 63 (44, 92) | 118 (82, 182) | <0.001 |
| VAT attenuation [HU] | -82 (-86, -76) | -87 (-90, -83) | <0.001 |
| VAT SD [HU] | 56 (52, 64) | 57 (53, 67) | <0.001 |
| VAT volume index [cm^3^/m^2^] | 294 (195, 425) | 462 (338, 624) | <0.001 |
| SAT attenuation [HU] | -99 (-103, -94) | -102 (-105, -99) | <0.001 |
| SAT SD [HU] | 37 (30, 45) | 44 (32, 56) | <0.001 |
| SAT volume index [cm^3^/m^2^] | 756 (502, 1,110) | 1,664 (1,181, 2,261) | <0.001 |
| SM attenuation [HU] | 32 (27, 37) | 28 (23, 33) | <0.001 |
| SM SD [HU] | 48 (39, 57) | 54 (42, 68) | <0.001 |
| SM volume index [cm^3^/m^2^] | 728 (599, 889) | 855 (711, 1,036) | <0.001 |
| Ratio SM/ATs volume | 0.61 (0.41, 0.88) | 0.35 (0.26, 0.50) | <0.001 |

Values are presented as N (%) or median (IQ1, IQ3)

AU – Agatston unit, ATs – adipose tissues, BMI – body mass index, CAC – coronary artery calcium, CAD – coronary artery disease, CT – computed tomography, EAT – epicardial adipose tissue, HU – Hounsfield unit, IMAT – intramuscular adipose tissue, LVEF – left ventricular ejection fraction, MPI – myocardial perfusion imaging, Ratio SM/ATs volume – ratio of skeletal muscle volume to all adipose tissues volume, SAT – subcutaneous adipose tissue, SD – standard deviation, SM – skeletal muscle, TPD – total perfusion deficit, VAT – visceral adipose tissue

**Supplementary Table 5. Youden index-based optimal cutoffs for our proposed volumetric body composition quantifications for all patients.**

|  | **Area under ROC [95%CI]** | **Youden index-based cutoff** |
| --- | --- | --- |
| Bone attenuation [HU] | 0.59 [0.57, 0.62] | 250.42 |
| Bone SD [HU] | 0.56 [0.54, 0.58] | 189.06 |
| Bone volume index [cm^3^/m^2^] | 0.58 [0.55, 0.6] | 306.98 |
| EAT attenuation [HU] | 0.55 [0.53, 0.58] | -63.08 |
| EAT SD [HU] | 0.54 [0.52, 0.56] | 48.08 |
| EAT volume index [cm^3^/m^2^] | 0.56 [0.53, 0.58] | 51.42 |
| IMAT attenuation [HU] | 0.61 [0.59, 0.64] | -67.78 |
| IMAT SD [HU] | 0.5 [0.48, 0.53] | 29.80 |
| IMAT volume index [cm^3^/m^2^] | 0.53 [0.51, 0.56] | 75.73 |
| VAT attenuation [HU] | 0.64 [0.61, 0.66] | -80.14 |
| VAT SD [HU] | 0.53 [0.51, 0.56] | 56.45 |
| VAT volume index [cm^3^/m^2^] | 0.51 [0.48, 0.53] | 500.20 |
| SAT attenuation [HU] | 0.64 [0.62, 0.66] | -100.73 |
| SAT SD [HU] | 0.49 [0.47, 0.52] | 45.86 |
| SAT volume index [cm^3^/m^2^] | 0.57 [0.55, 0.59] | 1117.13 |
| SM attenuation [HU] | 0.58 [0.56, 0.6] | 30.79 |
| SM SD [HU] | 0.53 [0.5, 0.55] | 45.55 |
| SM volume index [cm^3^/m^2^] | 0.55 [0.53, 0.58] | 597.16 |
| Ratio SM/ATs volume | 0.52 [0.5, 0.54] | 0.69 |

ATs – adipose tissues, CI – confidence interval, EAT – epicardial adipose tissue, HU – Hounsfield unit, IMAT – intramuscular adipose tissue, Ratio SM/ATs volume – ratio of skeletal muscle volume to all adipose tissues volume, ROC – receiver operating characteristic, SAT – subcutaneous adipose tissue, SD – standard deviation, SM – skeletal muscle, VAT – visceral adipose tissue

**Supplementary Table 6. Comparison between unadjusted and adjusted hazard ratios (HRs).** Unadjusted HRs were calculated from a Cox regression model over each factor independently. Adjusted HRs were calculated from multivariate Cox regression model over 11 clinical and perfusion factors and all the other body composition quantifications. Cutoffs from Supplementary Table 5 were used to categorize patients into low quantification group and high quantification group. Groups with low quantifications were used as reference group. For gender, female group was used as reference group. The HRs associated with age, body mass index (BMI), and log(CAC score+1) were for per unit increase. For diabetes mellitus, dyslipidemia, family history, hypertension, and smoking, patients without them were the reference groups. For left ventricular ejection fraction (LVEF), patients with LEVF ≥ 50% were considered high and used as the reference group. For stress total perfusion deficit (TPD), patients with stress TPD < 5% were considered low and used as the reference group.

| **Risk factors** | | **HR [95% CI], p-value** | | |
| --- | --- | --- | --- | --- |
|  |  | **Univariate** | **Multivariate adjusted for 11 clinical and imaging variables** | **Multivariable (adjusted for 11 clinical and imaging variables and other 18 body composition measures)** |
| 11 existing clinical and imaging risk factors | Age | 1.04 [1.04, 1.05], <0.0001 | – | 1.01 [1.01, 1.02], 0.0017 |
|  | BMI | 0.95 [0.94, 0.97], <0.0001 | – | 0.98 [0.96, 1], 0.0278 |
|  | Diabetes mellitus | 1.44 [1.22, 1.69], <0.0001 | – | 1.33 [1.12, 1.59], 0.0014 |
|  | Dyslipidemia | 1.1 [0.94, 1.29], 0.252 | – | 0.88 [0.74, 1.05], 0.1508 |
|  | Family history | 0.82 [0.68, 1], 0.0539 | – | 0.73 [0.6, 0.91], 0.0039 |
|  | Gender | 1.41 [1.19, 1.66], <0.0001 | – | 1.34 [1.05, 1.73], 0.0211 |
|  | Hypertension | 1.22 [1.03, 1.44], 0.0188 | – | 1.11 [0.93, 1.33], 0.2478 |
|  | log(CAC score+1) | 1.51 [1.41, 1.61], <0.0001 | – | 1.16 [1.07, 1.25], 0.0002 |
|  | LVEF | 2.8 [2.37, 3.31], <0.0001 | – | 1.5 [1.24, 1.81], <0.0001 |
|  | Smoking | 1.28 [1.04, 1.58], 0.0213 | – | 1.31 [1.05, 1.62], 0.0158 |
|  | Stress TPD | 1.62 [1.38, 1.9], <0.0001 | – | 1.28 [1.08, 1.52], 0.0049 |
| 19 volumetric body composition quantifications | Bone attenuation | **0.54 [0.46, 0.64], <0.0001** | **0.67 [0.57, 0.79], <0.0001** | **0.77 [0.62, 0.95], 0.0159** |
|  | Bone SD | **0.79 [0.67, 0.93], 0.0039** | **0.83 [0.7, 0.97], 0.0213** | 1.1 [0.89, 1.36], 0.3911 |
|  | Bone volume index | **1.34 [1.14, 1.59], 0.0005** | 1.04 [0.85, 1.29], 0.6835 | 1.16 [0.93, 1.46], 0.1922 |
|  | EAT attenuation | **1.85 [1.55, 2.2], <0.0001** | **2.02 [1.68, 2.43], <0.0001** | **1.55 [1.26, 1.9], <0.0001** |
|  | EAT SD | **1.62 [1.37, 1.9], <0.0001** | **1.58 [1.33, 1.88], <0.0001** | **1.5 [1.23, 1.83], 0.0001** |
|  | EAT volume index | **1.7 [1.44, 1.99], <0.0001** | **1.65 [1.39, 1.96], <0.0001** | **1.56 [1.28, 1.91], <0.0001** |
|  | IMAT attenuation | **2.17 [1.85, 2.54], <0.0001** | **1.68 [1.4, 2.01], <0.0001** | **1.3 [1.06, 1.6], 0.0124** |
|  | IMAT SD | **0.71 [0.54, 0.93], 0.0123** | 0.97 [0.72, 1.29], 0.8271 | 0.9 [0.65, 1.26], 0.5486 |
|  | IMAT volume index | **1.33 [1.12, 1.57], 0.0009** | **1.54 [1.27, 1.87], <0.0001** | **1.32 [1.05, 1.66], 0.0165** |
|  | VAT attenuation | **2.5 [2.13, 2.93], <0.0001** | **2.35 [1.96, 2.81], <0.0001** | **2.39 [1.92, 2.96], <0.0001** |
|  | VAT SD | 0.89 [0.76, 1.05], 0.1642 | 1.06 [0.9, 1.25], 0.4853 | **0.78 [0.63, 0.96], 0.0219** |
|  | VAT volume index | 1.12 [0.94, 1.32], 0.2072 | 1.06 [0.87, 1.29], 0.5804 | 1.04 [0.83, 1.31], 0.7274 |
|  | Ratio SM/AT | **1.24 [1.04, 1.49], 0.0153** | 1.06 [0.86, 1.31], 0.5820 | 1.15 [0.9, 1.48], 0.2671 |
|  | SAT attenuation | **2.1 [1.76, 2.49], <0.0001** | **1.53 [1.26, 1.85], <0.0001** | **1.3 [1.05, 1.61], 0.0145** |
|  | SAT SD | 1.07 [0.9, 1.28], 0.4458 | **1.3 [1.07, 1.57], 0.0070** | 1.01 [0.79, 1.29], 0.9502 |
|  | SAT volume index | **0.6 [0.51, 0.7], <0.0001** | 0.83 [0.66, 1.04], 0.1025 | 0.94 [0.74, 1.2], 0.6429 |
|  | SM attenuation | **0.61 [0.51, 0.72], <0.0001** | **0.58 [0.48, 0.7], <0.0001** | **0.54 [0.44, 0.67], <0.0001** |
|  | SM SD | **1.67 [1.41, 1.99], <0.0001** | **1.91 [1.59, 2.3], <0.0001** | **1.64 [1.27, 2.12], 0.0002** |
|  | SM volume index | **0.49 [0.41, 0.59], <0.0001** | **0.52 [0.42, 0.65], <0.0001** | **0.56 [0.44, 0.71], <0.0001** |

ATs – adipose tissues, CAC – coronary artery calcium, CI – confidence interval, CT – computed tomography, EAT – epicardial adipose tissue, HU – Hounsfield unit, IMAT – intramuscular adipose tissue, MPI – myocardial perfusion imaging, SAT – subcutaneous adipose tissue, SD – standard deviation, SM – skeletal muscle, Ratio SM/AT - ratio of skeletal muscle volume to all adipose tissues volume, VAT – visceral adipose tissue

**Supplementary Table 7. Comparison of unadjusted hazard ratios (HRs) between male and female patients.**

The cutoffs are from Supplementary Table 5.

| **Factor** | **Unadjusted hazard Ratios [95% CI], p-value** | |
| --- | --- | --- |
|  | **Male** | **Female** |
| Bone attenuation [HU] | **0.53 [0.43, 0.64], <0.0001** | **0.53 [0.41, 0.7], <0.0001** |
| Bone SD [HU] | **0.79 [0.65, 0.96], 0.0207** | 0.78 [0.6, 1.02], 0.0677 |
| Bone volume index [cm^3^/m^2^] | 1.09 [0.83, 1.44], 0.5239 | 1.24 [0.93, 1.63], 0.1381 |
| EAT attenuation [HU] | **2.01 [1.61, 2.52], <0.0001** | **1.61 [1.22, 2.12], 0.0008** |
| EAT SD [HU] | **1.86 [1.5, 2.29], <0.0001** | 1.23 [0.95, 1.6], 0.1197 |
| EAT volume index [cm^3^/m^2^] | **1.65 [1.35, 2.02], <0.0001** | **1.73 [1.33, 2.25], <0.0001** |
| IMAT attenuation [HU] | **2.03 [1.66, 2.48], <0.0001** | **2.64 [2.03, 3.45], <0.0001** |
| IMAT SD [HU] | 0.82 [0.57, 1.18], 0.2842 | **0.57 [0.38, 0.85], 0.0063** |
| IMAT volume index [cm^3^/m^2^] | **1.32 [1.06, 1.66], 0.0150** | 1.17 [0.9, 1.52], 0.2401 |
| VAT attenuation [HU] | **2.79 [2.28, 3.42], <0.0001** | **2.71 [2.08, 3.54], <0.0001** |
| VAT SD [HU] | 0.99 [0.81, 1.21], 0.9201 | 0.78 [0.6, 1.01], 0.0623 |
| VAT volume index [cm^3^/m^2^] | 0.96 [0.79, 1.18], 0.7227 | 1.13 [0.8, 1.6], 0.4884 |
| SAT attenuation [HU] | **1.37 [1.09, 1.72], 0.0069** | **3.22 [2.46, 4.21], <0.0001** |
| SAT SD [HU] | 1.14 [0.92, 1.41], 0.2346 | 0.88 [0.64, 1.2], 0.4085 |
| SAT volume index [cm^3^/m^2^] | **0.73 [0.58, 0.91], 0.0054** | **0.52 [0.4, 0.69], <0.0001** |
| SM attenuation [HU] | **0.47 [0.39, 0.58], <0.0001** | **0.54 [0.37, 0.78], 0.0010** |
| SM SD [HU] | **1.73 [1.38, 2.18], <0.0001** | **1.49 [1.14, 1.95], 0.0038** |
| SM volume index [cm^3^/m^2^] | **0.25 [0.18, 0.33], <0.0001** | **0.47 [0.36, 0.61], <0.0001** |
| Ratio SM/ATs volume | 0.97 [0.79, 1.2], 0.8065 | **1.89 [1.28, 2.79], 0.0014** |

ATs – adipose tissues, CI – confidence interval, EAT – epicardial adipose tissue, HU – Hounsfield unit, IMAT – intramuscular adipose tissue, Ratio SM/ATs volume – ratio of skeletal muscle volume to all adipose tissues volume, SAT – subcutaneous adipose tissue, SD – standard deviation, SM – skeletal muscle, VAT – visceral adipose tissue

**Supplementary Table 8. Comparison of adjusted hazard ratios (HRs, adjusted for 10 remaining clinical and imaging risk factors) between male and female patients**. The cutoffs are from Supplementary Table 5.

| **Factor** | **Adjusted hazard Ratios [95% CI], p-value** | |
| --- | --- | --- |
|  | **Male** | **Female** |
| Bone attenuation [HU] | **0.64 [0.52, 0.79], <0.0001** | 0.76 [0.57, 1], 0.0522 |
| Bone SD [HU] | **0.79 [0.64, 0.96], 0.0207** | 0.92 [0.7, 1.2], 0.5227 |
| Bone volume index [cm^3^/m^2^] | 0.96 [0.72, 1.28], 0.7938 | 1.08 [0.8, 1.46], 0.5996 |
| EAT attenuation [HU] | **2.28 [1.8, 2.89], <0.0001** | **1.66 [1.24, 2.23], 0.0007** |
| EAT SD [HU] | **1.78 [1.43, 2.23], <0.0001** | **1.35 [1.02, 1.79], 0.0334** |
| EAT volume index [cm^3^/m^2^] | **1.64 [1.31, 2.05], <0.0001** | **1.66 [1.26, 2.19], 0.0003** |
| IMAT attenuation [HU] | **1.62 [1.28, 2.04], <0.0001** | **1.79 [1.34, 2.41], 0.0001** |
| IMAT SD [HU] | 1.03 [0.7, 1.52], 0.8682 | 0.92 [0.59, 1.44], 0.7255 |
| IMAT volume index [cm^3^/m^2^] | **1.5 [1.16, 1.93], 0.0017** | **1.64 [1.21, 2.23], 0.0016** |
| VAT attenuation [HU] | **2.42 [1.94, 3.04], <0.0001** | **2.28 [1.7, 3.07], <0.0001** |
| VAT SD [HU] | 1.12 [0.91, 1.38], 0.2986 | 1 [0.76, 1.31], 0.9923 |
| VAT volume index [cm^3^/m^2^] | 0.98 [0.78, 1.23], 0.8541 | 1.27 [0.87, 1.85], 0.2079 |
| SAT attenuation [HU] | 1.06 [0.83, 1.35], 0.6314 | **2.36 [1.77, 3.15], <0.0001** |
| SAT SD [HU] | **1.33 [1.05, 1.67], 0.016** | 1.25 [0.9, 1.76], 0.1866 |
| SAT volume index [cm^3^/m^2^] | 0.91 [0.68, 1.2], 0.5 | 0.72 [0.5, 1.02], 0.0666 |
| SM attenuation [HU] | **0.58 [0.46, 0.72], <0.0001** | **0.61 [0.41, 0.9], 0.0119** |
| SM SD [HU] | **1.99 [1.56, 2.54], <0.0001** | **1.9 [1.43, 2.52], <0.0001** |
| SM volume index [cm^3^/m^2^] | **0.37 [0.27, 0.5], <0.0001** | **0.66 [0.49, 0.88], 0.0049** |
| Ratio SM/ATs volume | 0.97 [0.76, 1.23], 0.7976 | **1.6 [1.03, 2.48], 0.0350** |

ATs – adipose tissues, CI – confidence interval, EAT – epicardial adipose tissue, HU – Hounsfield unit, IMAT – intramuscular adipose tissue, Ratio SM/ATs volume – ratio of skeletal muscle volume to all adipose tissues volume, SAT – subcutaneous adipose tissue, SD – standard deviation, SM – skeletal muscle, VAT – visceral adipose tissue

**Supplementary Table 9. Comparison of adjusted hazard ratios (HRs, adjusted for 10 remaining clinical and imaging risk factors and 18 remaining body composition risk factors) between male and female patients.** The cutoffs are from Supplementary Table 5.

| **Factor** | **Adjusted hazard Ratios [95% CI], p-value** | |
| --- | --- | --- |
|  | **Male** | **Female** |
| Bone attenuation [HU] | 0.78 [0.59, 1.02], 0.0705 | 0.82 [0.57, 1.17], 0.2751 |
| Bone SD [HU] | 1.04 [0.79, 1.38], 0.7551 | 1.21 [0.86, 1.7], 0.2759 |
| Bone volume index [cm^3^/m^2^] | 1.3 [0.94, 1.81], 0.1108 | 1 [0.72, 1.39], 0.9978 |
| EAT attenuation [HU] | **1.76 [1.35, 2.31], <0.0001** | 1.24 [0.89, 1.71], 0.2043 |
| EAT SD [HU] | **1.76 [1.36, 2.27], <0.0001** | 1.26 [0.91, 1.73], 0.1642 |
| EAT volume index [cm^3^/m^2^] | **1.43 [1.1, 1.87], 0.0086** | **1.69 [1.23, 2.31], 0.0012** |
| IMAT attenuation [HU] | 1.25 [0.94, 1.66], 0.1210 | 1.23 [0.89, 1.71], 0.2061 |
| IMAT SD [HU] | 0.94 [0.6, 1.47], 0.7885 | 0.97 [0.58, 1.63], 0.9225 |
| IMAT volume index [cm^3^/m^2^] | 1.31 [0.97, 1.77], 0.0734 | 1.29 [0.89, 1.85], 0.1795 |
| VAT attenuation [HU] | **2.43 [1.82, 3.24], <0.0001** | **2.4 [1.68, 3.43], <0.0001** |
| VAT SD [HU] | **0.73 [0.55, 0.97], 0.0290** | 0.8 [0.57, 1.13], 0.2031 |
| VAT volume index [cm^3^/m^2^] | 0.94 [0.7, 1.24], 0.6482 | 1.28 [0.84, 1.94], 0.2445 |
| SAT attenuation [HU] | 0.98 [0.74, 1.3], 0.9016 | **1.85 [1.32, 2.59], 0.0004** |
| SAT SD [HU] | 1.08 [0.79, 1.47], 0.6419 | 0.8 [0.52, 1.22], 0.3021 |
| SAT volume index [cm^3^/m^2^] | 0.97 [0.71, 1.32], 0.8387 | 0.98 [0.65, 1.48], 0.9249 |
| SM attenuation [HU] | **0.53 [0.41, 0.69], <0.0001** | **0.58 [0.38, 0.88], 0.0107** |
| SM SD [HU] | **1.55 [1.09, 2.2], 0.0146** | **1.92 [1.3, 2.83], 0.0010** |
| SM volume index [cm^3^/m^2^] | **0.45 [0.31, 0.64], <0.0001** | **0.6 [0.43, 0.83], 0.0023** |
| Ratio SM/ATs volume | 1.09 [0.81, 1.48], 0.5559 | **1.68 [1.01, 2.8], 0.0470** |

ATs – adipose tissues, CI – confidence interval, EAT – epicardial adipose tissue, HU – Hounsfield unit, IMAT – intramuscular adipose tissue, Ratio SM/ATs volume – ratio of skeletal muscle volume to all adipose tissues volume, SAT – subcutaneous adipose tissue, SD – standard deviation, SM – skeletal muscle, VAT – visceral adipose tissue

**Supplementary Table 10. Comparison of unadjusted hazard ratios among Black or African America and White patients.** The cutoffs are from Supplementary Table 5.

| **Factor** | **Hazard Ratios [95% CI], p-value** | |
| --- | --- | --- |
|  | **Black or African American** | **White** |
| Bone attenuation [HU] | 0.6 [0.36, 1], 0.0518 | **0.59 [0.45, 0.79], 0.0004** |
| Bone SD [HU] | **0.56 [0.35, 0.9], 0.0168** | 1.21 [0.89, 1.66], 0.2212 |
| Bone volume index [cm^3^/m^2^] | **1.67 [1.06, 2.63], 0.027** | 1.18 [0.88, 1.59], 0.2680 |
| EAT attenuation [HU] | 1.53 [0.93, 2.51], 0.0912 | **2.08 [1.53, 2.82], <0.0001** |
| EAT SD [HU] | **2.05 [1.28, 3.29], 0.0030** | **1.37 [1.02, 1.85], 0.0388** |
| EAT volume index [cm^3^/m^2^] | 1.55 [0.92, 2.62], 0.1014 | **1.5 [1.12, 2.01], 0.0064** |
| IMAT attenuation [HU] | **1.8 [1.15, 2.83], 0.0100** | **2.12 [1.6, 2.82], <0.0001** |
| IMAT SD [HU] | 0.94 [0.43, 2.05], 0.8767 | 0.96 [0.57, 1.64], 0.8869 |
| IMAT volume index [cm^3^/m^2^] | 1.36 [0.85, 2.18], 0.2054 | **1.38 [1.02, 1.87], 0.0363** |
| VAT attenuation [HU] | **1.99 [1.26, 3.15], 0.0032** | **2.91 [2.19, 3.87], <0.0001** |
| VAT SD [HU] | 1.75 [0.92, 3.34], 0.0893 | 0.98 [0.72, 1.33], 0.8905 |
| VAT volume index [cm^3^/m^2^] | 1.63 [0.84, 3.18], 0.1491 | 0.82 [0.59, 1.12], 0.2160 |
| SAT attenuation [HU] | **2.31 [1.42, 3.74], 0.0007** | **1.77 [1.33, 2.37], 0.0001** |
| SAT SD [HU] | 1.39 [0.85, 2.27], 0.1851 | 1.25 [0.92, 1.69], 0.1536 |
| SAT volume index [cm^3^/m^2^] | **0.59 [0.37, 0.92], 0.0204** | **0.64 [0.48, 0.85], 0.0025** |
| SM attenuation [HU] | 0.65 [0.41, 1.05], 0.0774 | **0.55 [0.4, 0.75], 0.0002** |
| SM SD [HU] | **1.81 [1.09, 3.01], 0.0221** | **1.62 [1.16, 2.25], 0.0044** |
| SM volume index [cm^3^/m^2^] | **0.46 [0.26, 0.82], 0.0088** | **0.45 [0.33, 0.61], <0.0001** |
| Ratio SM/ATs volume | 1.42 [0.89, 2.29], 0.1429 | 1 [0.71, 1.39], 0.9925 |

ATs – adipose tissues, CI – confidence interval, EAT – epicardial adipose tissue, HU – Hounsfield unit, IMAT – intramuscular adipose tissue, Ratio SM/ATs volume – ratio of skeletal muscle volume to all adipose tissues volume, SAT – subcutaneous adipose tissue, SD – standard deviation, SM – skeletal muscle, VAT – visceral adipose tissue

**Supplementary Table 11. Comparison of unadjusted hazard ratios (HRs) between patients with <65 and ≥65 years.** The cutoffs are from Supplementary Table 5.

| **Factor** | **Hazard Ratios [95% CI], p-value** | |
| --- | --- | --- |
|  | **<65 years** | **≥65 years** |
| Bone attenuation [HU] | **0.6 [0.45, 0.81], 0.0008** | **0.68 [0.56, 0.83], 0.0004** |
| Bone SD [HU] | **0.63 [0.47, 0.84], 0.0016** | 1.02 [0.85, 1.23], 0.8194 |
| Bone volume index [cm^3^/m^2^] | **1.55 [1.14, 2.1], 0.0047** | 1.2 [0.99, 1.46], 0.0632 |
| EAT attenuation [HU] | **1.51 [1.1, 2.07], 0.0113** | **2.16 [1.76, 2.65], <0.0001** |
| EAT SD [HU] | **1.56 [1.15, 2.13], 0.0046** | **1.76 [1.45, 2.15], <0.0001** |
| EAT volume index [cm^3^/m^2^] | **1.43 [1.05, 1.94], 0.0214** | **1.52 [1.26, 1.83], <0.0001** |
| IMAT attenuation [HU] | **2.48 [1.86, 3.33], <0.0001** | **1.69 [1.4, 2.03], <0.0001** |
| IMAT SD [HU] | 1.08 [0.79, 1.47], 0.6349 | **1.57 [1.29, 1.93], <0.0001** |
| IMAT volume index [cm^3^/m^2^] | **1.44 [1.05, 1.96], 0.0219** | **1.3 [1.07, 1.58], 0.0072** |
| VAT attenuation [HU] | **2.97 [2.22, 3.97], <0.0001** | **2.22 [1.85, 2.67], <0.0001** |
| VAT SD [HU] | 0.93 [0.69, 1.25], 0.6215 | 1.06 [0.88, 1.28], 0.5211 |
| VAT volume index [cm^3^/m^2^] | 0.93 [0.67, 1.3], 0.6832 | 1.06 [0.88, 1.28], 0.5485 |
| SAT attenuation [HU] | **2.25 [1.65, 3.08], <0.0001** | **1.86 [1.53, 2.27], <0.0001** |
| SAT SD [HU] | 1.09 [0.8, 1.49], 0.5739 | 1.2 [0.98, 1.47], 0.0769 |
| SAT volume index [cm^3^/m^2^] | **0.59 [0.44, 0.79], 0.0004** | **0.64 [0.53, 0.77], <0.0001** |
| SM attenuation [HU] | **0.62 [0.46, 0.84], 0.0016** | **0.75 [0.61, 0.91], 0.0046** |
| SM SD [HU] | **1.53 [1.11, 2.1], 0.0091** | **1.82 [1.49, 2.23], <0.0001** |
| SM volume index [cm^3^/m^2^] | **0.52 [0.35, 0.77], 0.0012** | **0.62 [0.5, 0.76], <0.0001** |
| Ratio SM/ATs volume | 1.44 [0.91, 2.27], 0.1181 | **1.5 [1.15, 1.95], 0.0027** |

ATs – adipose tissues, CI – confidence interval, EAT – epicardial adipose tissue, HU – Hounsfield unit, IMAT – intramuscular adipose tissue, Ratio SM/ATs volume – ratio of skeletal muscle volume to all adipose tissues volume, SAT – subcutaneous adipose tissue, SD – standard deviation, SM – skeletal muscle, VAT – visceral adipose tissue

**Supplementary Table 12. Comparison of unadjusted hazard ratios (HRs) between patients with BMI <30 kg/m^2^ and BMI ≥30 kg/m^2^.** The cutoffs are from Supplementary Table 5.

| **Factor** | **Hazard Ratios [95% CI], p-value** | |
| --- | --- | --- |
|  | **BMI <30 kg/m^2^** | **BMI ≥30 kg/m^2^** |
| Bone attenuation [HU] | **0.52 [0.42, 0.64], <0.0001** | **0.58 [0.45, 0.74], <0.0001** |
| Bone SD [HU] | **0.75 [0.61, 0.92], 0.0065** | 0.85 [0.66, 1.09], 0.2046 |
| Bone volume index [cm^3^/m^2^] | **1.26 [1.02, 1.55], 0.0291** | **1.73 [1.3, 2.31], 0.0002** |
| EAT attenuation [HU] | **1.65 [1.3, 2.08], <0.0001** | **1.87 [1.43, 2.44], <0.0001** |
| EAT SD [HU] | **1.67 [1.36, 2.05], <0.0001** | **2.31 [1.73, 3.09], <0.0001** |
| EAT volume index [cm^3^/m^2^] | **1.78 [1.44, 2.19], <0.0001** | **2.36 [1.78, 3.13], <0.0001** |
| IMAT attenuation [HU] | **2.25 [1.79, 2.82], <0.0001** | **1.56 [1.16, 2.1], 0.0032** |
| IMAT SD [HU] | 0.91 [0.69, 1.22], 0.5418 | 0.4 [0.13, 1.24], 0.1103 |
| IMAT volume index [cm^3^/m^2^] | **1.65 [1.35, 2.03], <0.0001** | **2.38 [1.58, 3.58], <0.0001** |
| VAT attenuation [HU] | **2.33 [1.89, 2.88], <0.0001** | **2.24 [1.70, 2.97], <0.0001** |
| VAT SD [HU] | **0.78 [0.63, 0.97], 0.0251** | 1.19 [0.92, 1.54], 0.1749 |
| VAT volume index [cm^3^/m^2^] | 1.17 [0.9, 1.53], 0.2355 | **1.56 [1.21, 2.01], 0.0006** |
| SAT attenuation [HU] | **1.9 [1.48, 2.44], <0.0001** | **1.94 [1.51, 2.5], <0.0001** |
| SAT SD [HU] | 1.07 [0.81, 1.4], 0.6391 | **1.5 [1.16, 1.95], 0.0021** |
| SAT volume index [cm^3^/m^2^] | **0.65 [0.5, 0.84], 0.0012** | 0.81 [0.6, 1.09], 0.1624 |
| SM attenuation [HU] | **0.48 [0.39, 0.59], <0.0001** | **0.61 [0.45, 0.82], 0.0012** |
| SM SD [HU] | **1.47 [1.18, 1.83], 0.0005** | **2.47 [1.82, 3.35], <0.0001** |
| SM volume index [cm^3^/m^2^] | **0.52 [0.41, 0.64], <0.0001** | **0.64 [0.44, 0.94], 0.0222** |
| Ratio SM/ATs volume | 1.02 [0.83, 1.26], 0.851 | 0.92 [0.57, 1.49], 0.7298 |

ATs – adipose tissues, BMI – body mass index, CI – confidence interval, EAT – epicardial adipose tissue, HU – Hounsfield unit, IMAT – intramuscular adipose tissue, Ratio SM/ATs volume – ratio of skeletal muscle volume to all adipose tissues volume, SAT – subcutaneous adipose tissue, SD – standard deviation, SM – skeletal muscle, VAT – visceral adipose tissue

**Supplementary Figures**

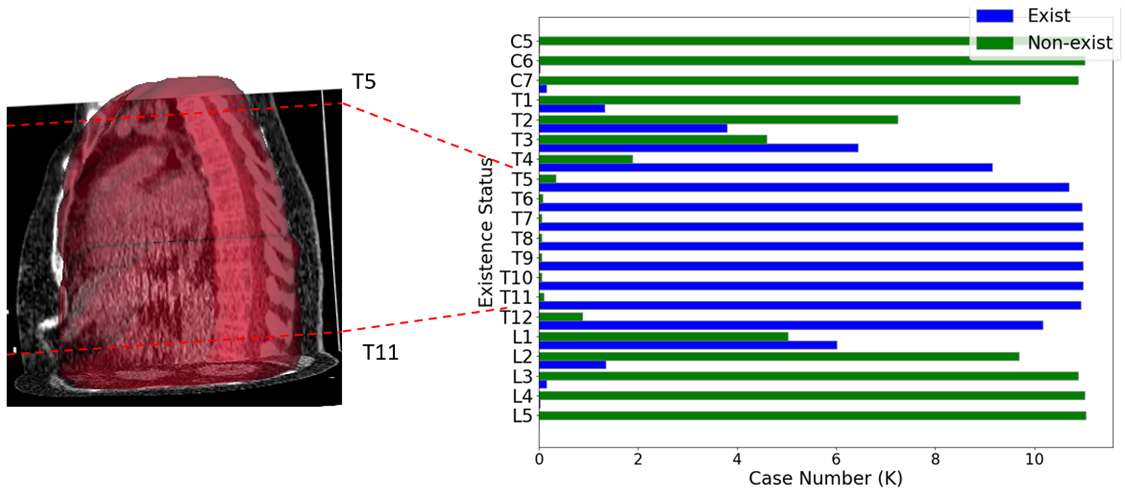

**Supplementary Figure 1. Histogram of visible vertebrae successfully, and automatically segmented from computed tomography attenuation correction maps in patients undergoing single-photon emission computed tomography (SPECT)/CT myocardial perfusion imaging.** Vertebrae included in the model (T5-T11) were visible in 96.4% of CT attenuation correction scans. Volumetric body composition quantification measures are computed from the sub-volume T5-T11.

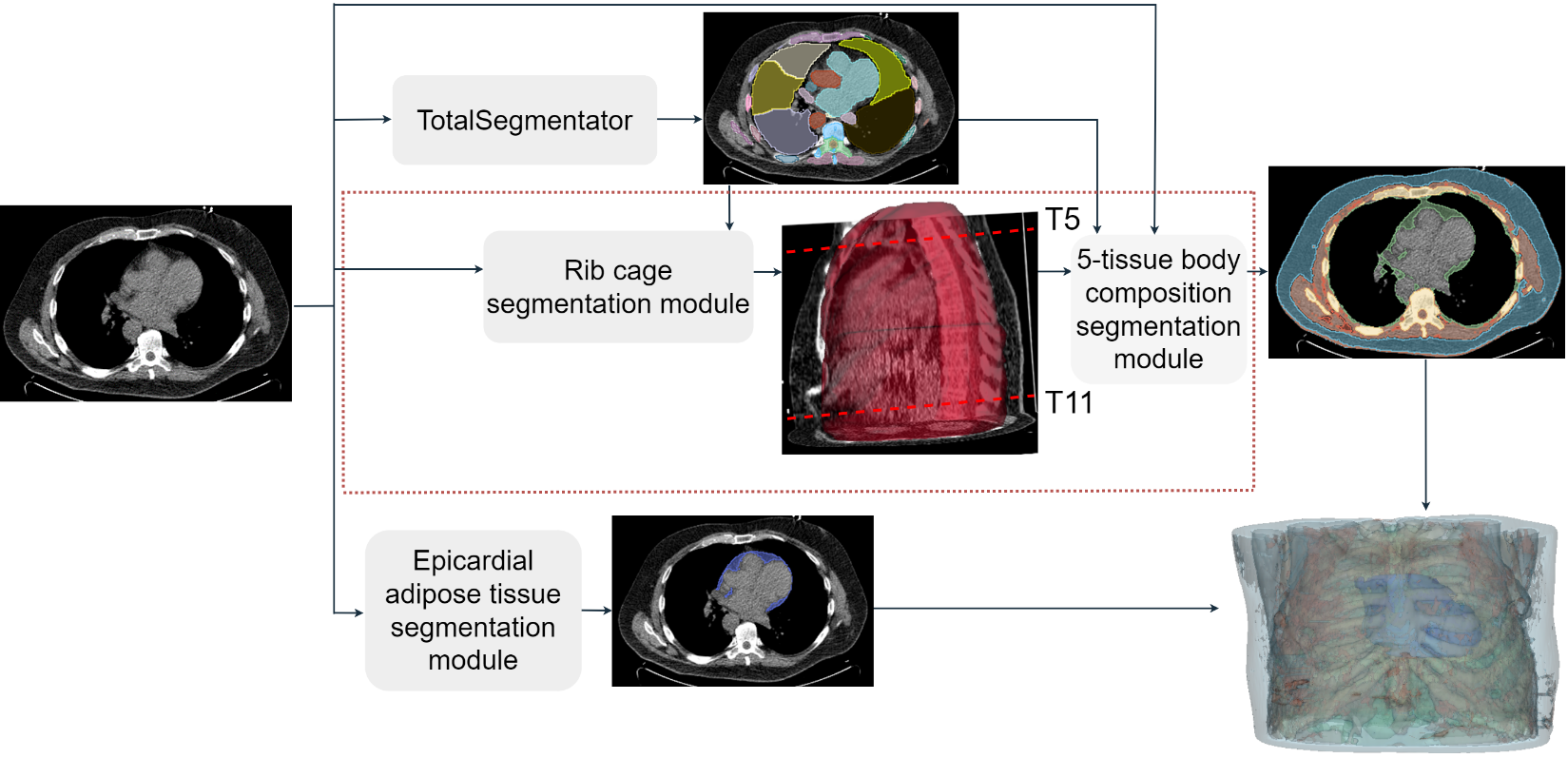

**Supplementary Figure 2. Pipeline of our proposed image processing-based body composition segmentation framework.** The proposed framework included our proposed body composition segmentation approach (the modules in the dashed box), a publicly available and trained TotalSegmentator v2 model (the TS v2 module), a previously trained epicardial adipose tissue (EAT) segmentation model (the epicardial adipose tissue segmentation module), and the body segmentation module (the Body module).^1-3^ The proposed body composition segmentation approach was based on the trained TotalSegmentator v2 and the body segmentation modules. The key idea was to first separate the body parts inside the rib cage from those outside the rib cage, and then apply thresholding to distinguish the bone, the muscle, and the fat which had very different Hounsfield Unit (HU) ranges. Our proposed body composition segmentation approach consisted of two core modules with the image processing-based rib cage segmentation module (primarily the convex hull operations over all foreground segmentations inside the rib cage) for segmenting the rib cage, and the body composition segmentation module (primarily the thresholding operations) for segmenting five body compositions (the bone, the skeletal muscle, the subcutaneous adipose tissue, the intramuscular adipose tissue, and the visceral adipose tissue (VAT)). By combining the EAT mask from the trained EAT segmentation model, we could further segment the EAT and non-EAT VAT to get the segmentation of six tissues.

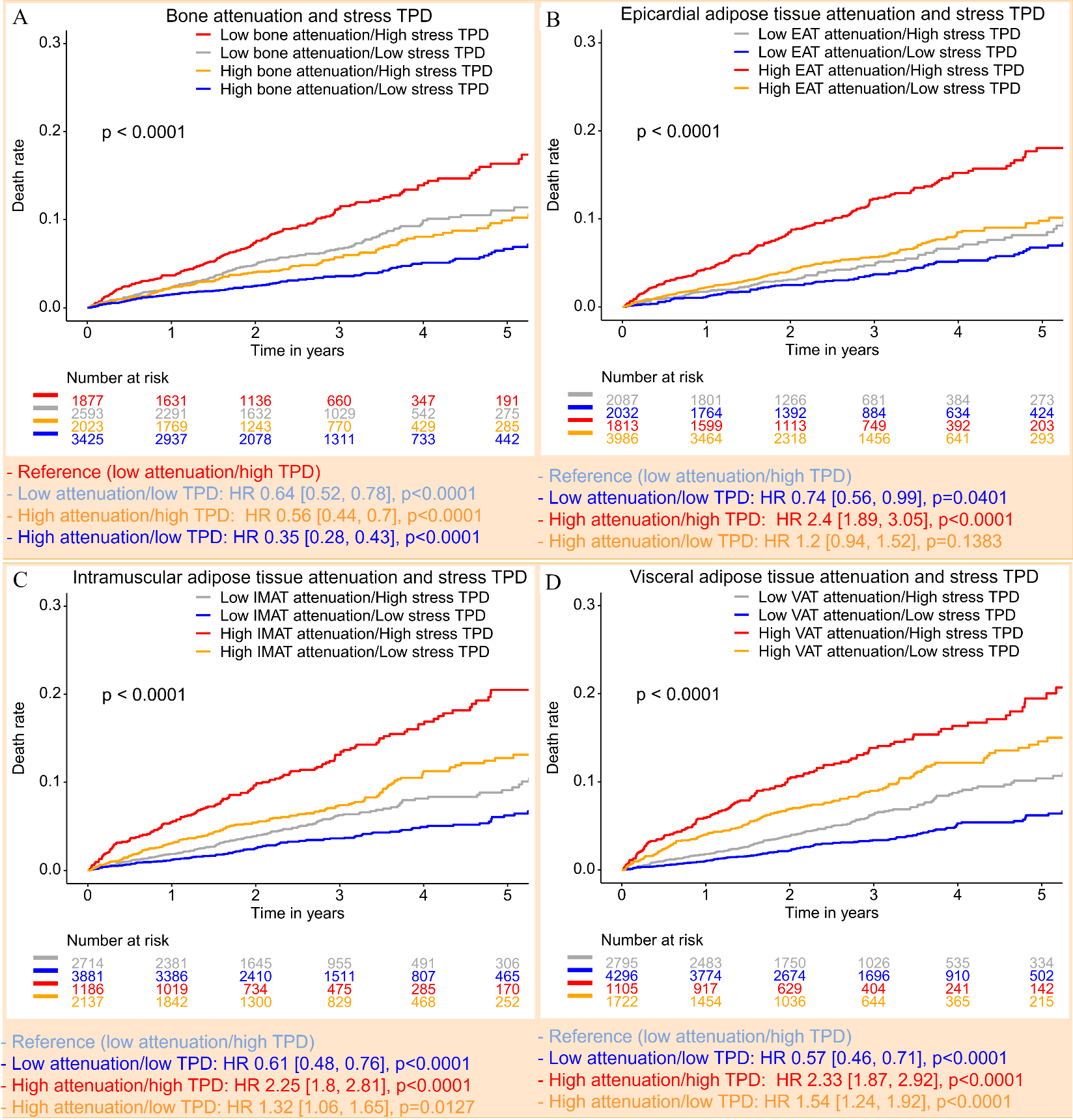

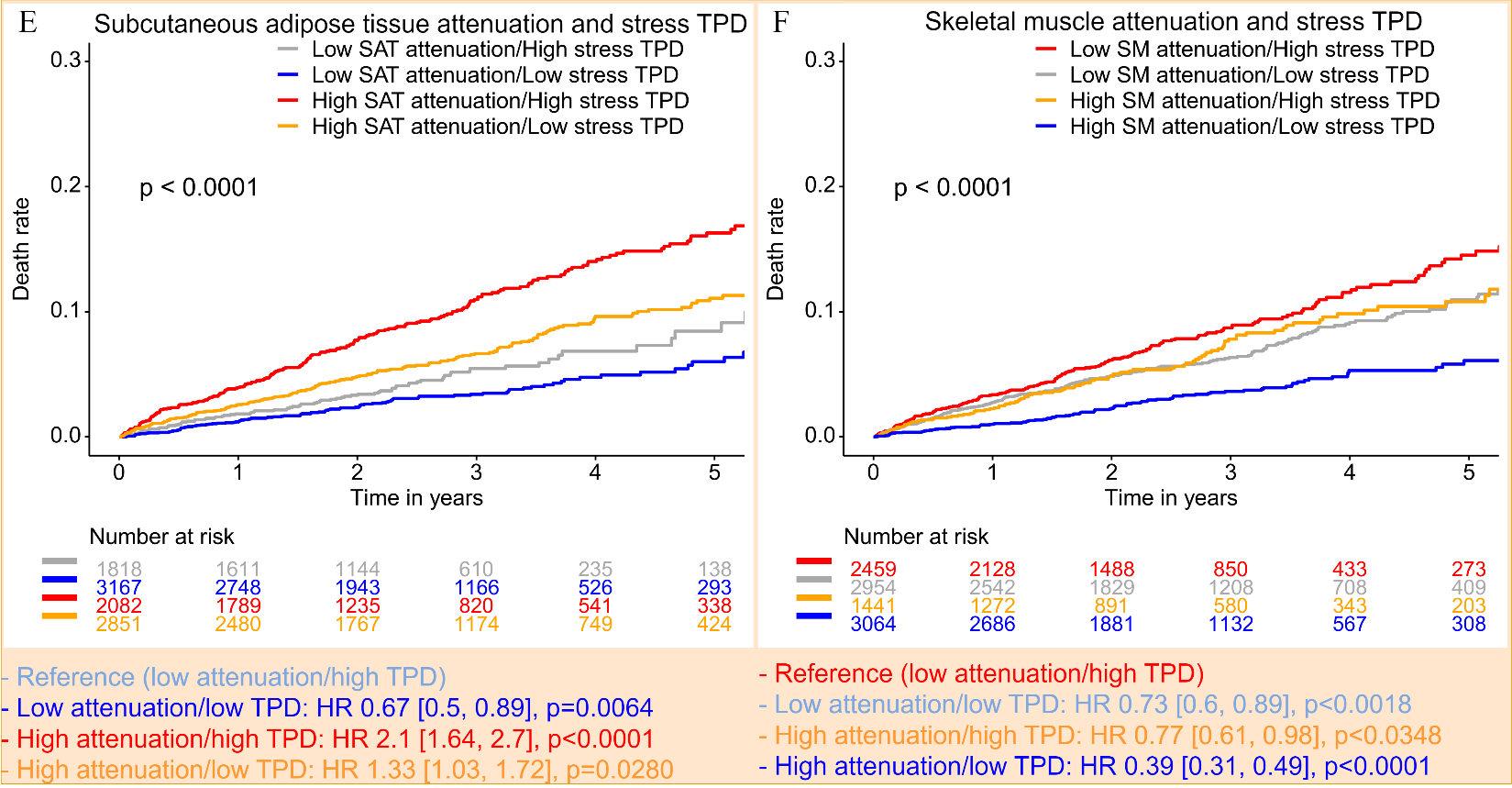

**Supplementary Figure 3. Kaplan-Meier curves for risk stratification with volumetric body composition attenuation and stress total perfusion deficit (TPD) in all patients.** **A:** bone, **B:** EAT – epicardial adipose tissue, **C:** IMAT – intramuscular adipose tissue, **D:** VAT – visceral adipose tissue, **E:** SAT – subcutaneous adipose tissue, **F:** SM – skeletal muscle. The cutoffs were from Suppl. Table 5. HR – hazard ratio

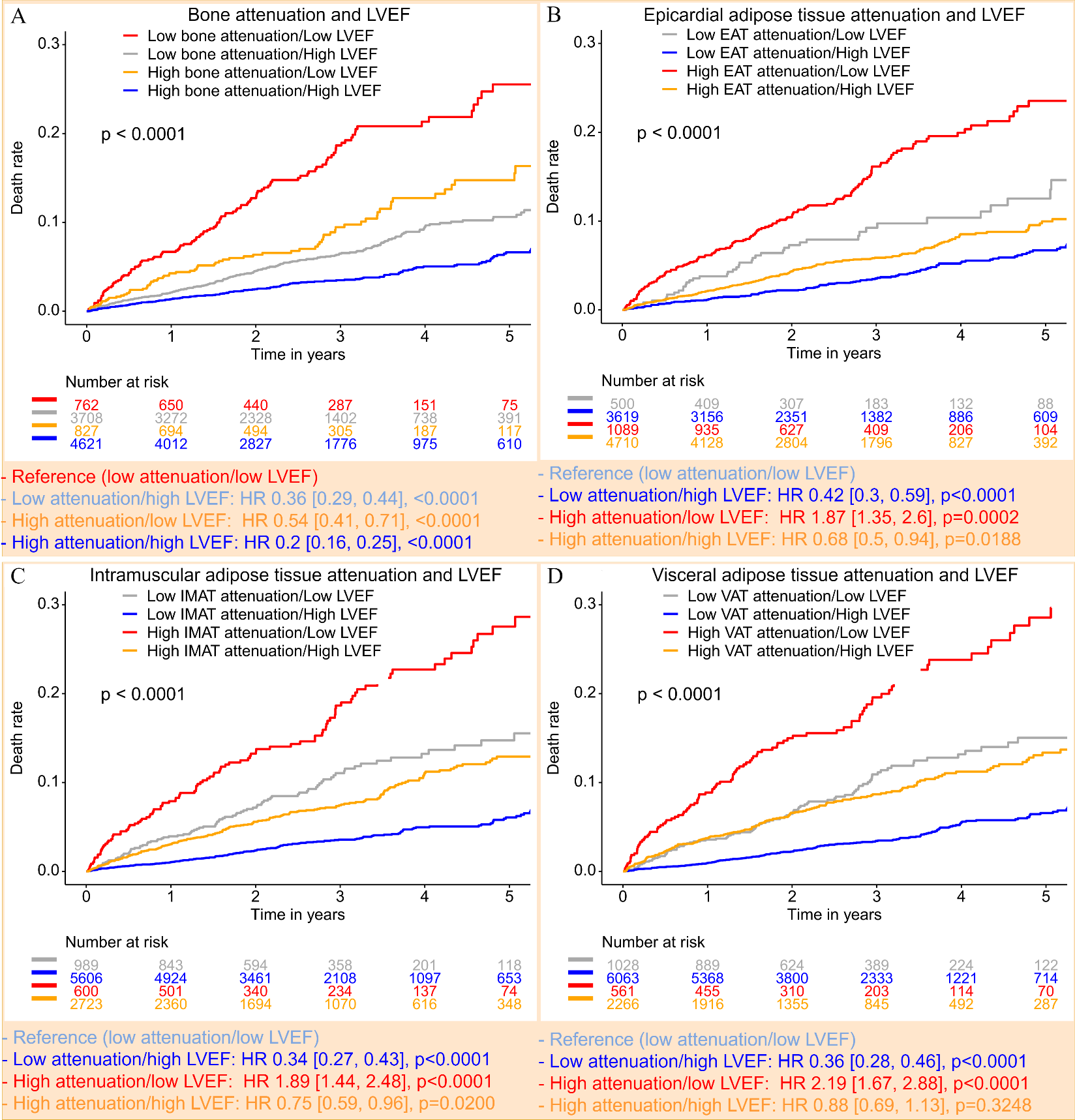

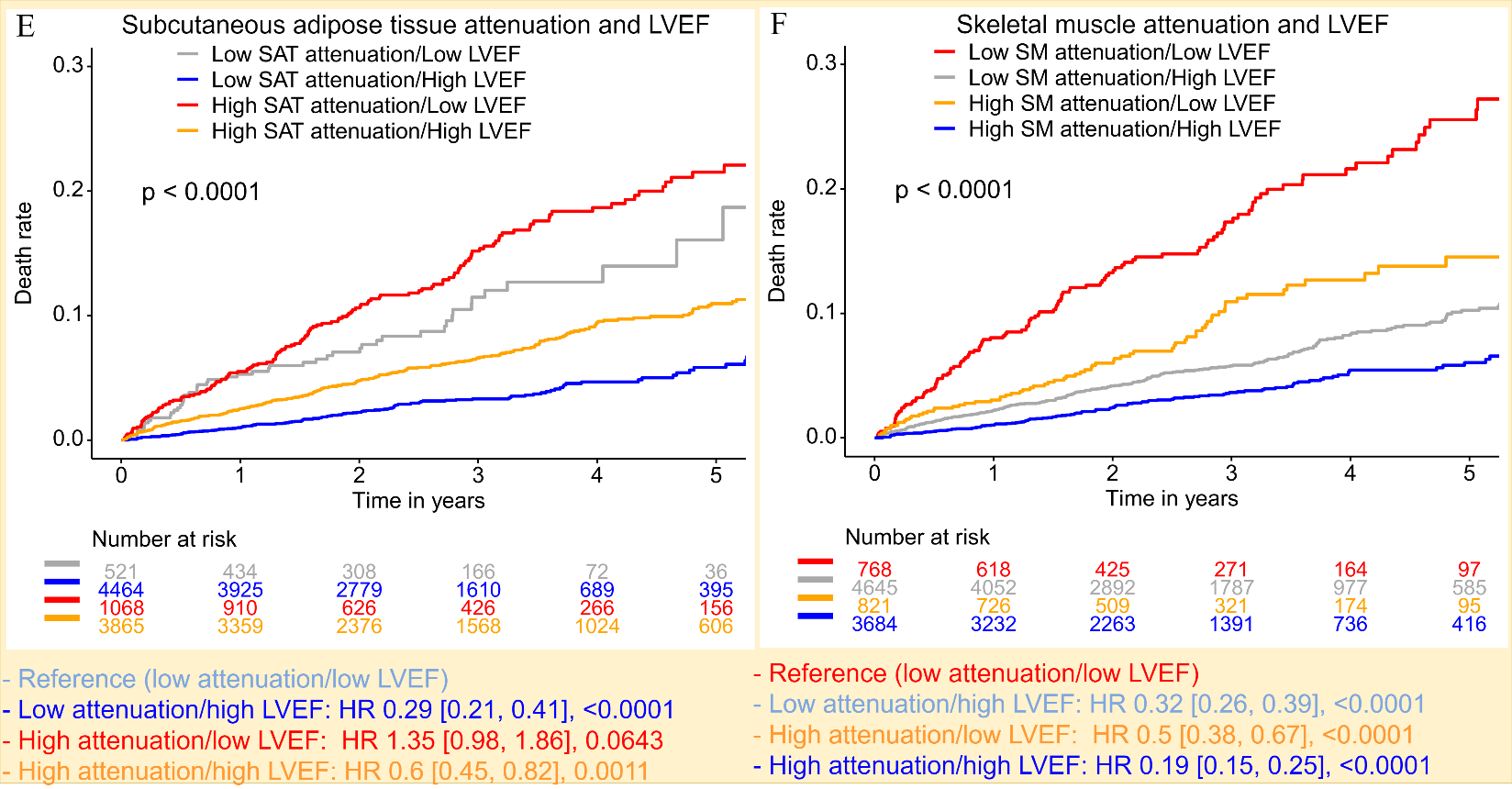

**Supplementary Figure 4. Kaplan-Meier curves for risk stratification with volumetric body composition attenuation and left ventricular ejection fraction (LVEF) in all patients.** **A:** bone, **B:** EAT – epicardial adipose tissue, **C:** IMAT – intramuscular adipose tissue, **D:** VAT – visceral adipose tissue, **E:** SAT – subcutaneous adipose tissue, **F:** SM – skeletal muscle. The cutoffs were from Suppl. Table 5. HR – hazard ratio.

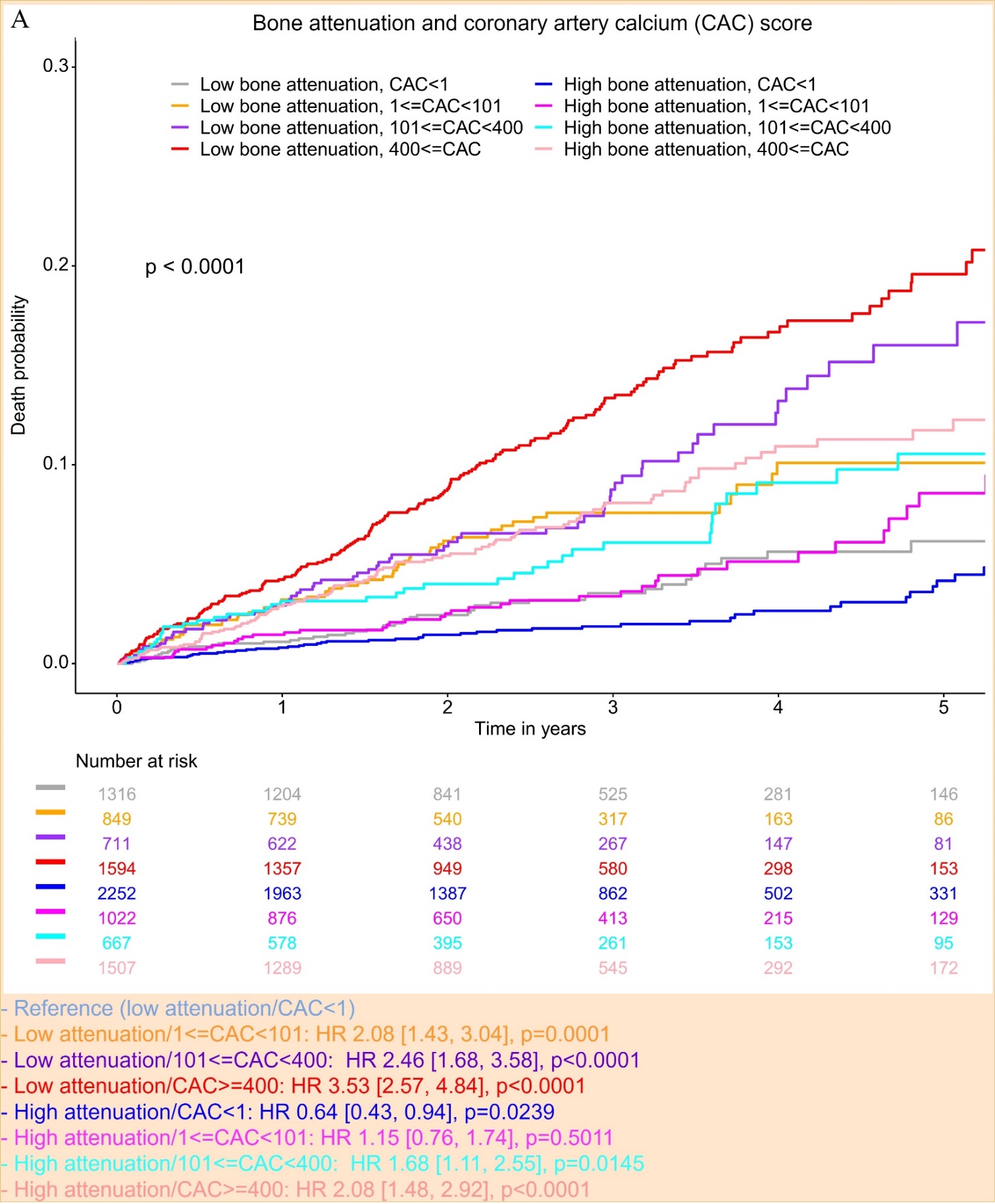

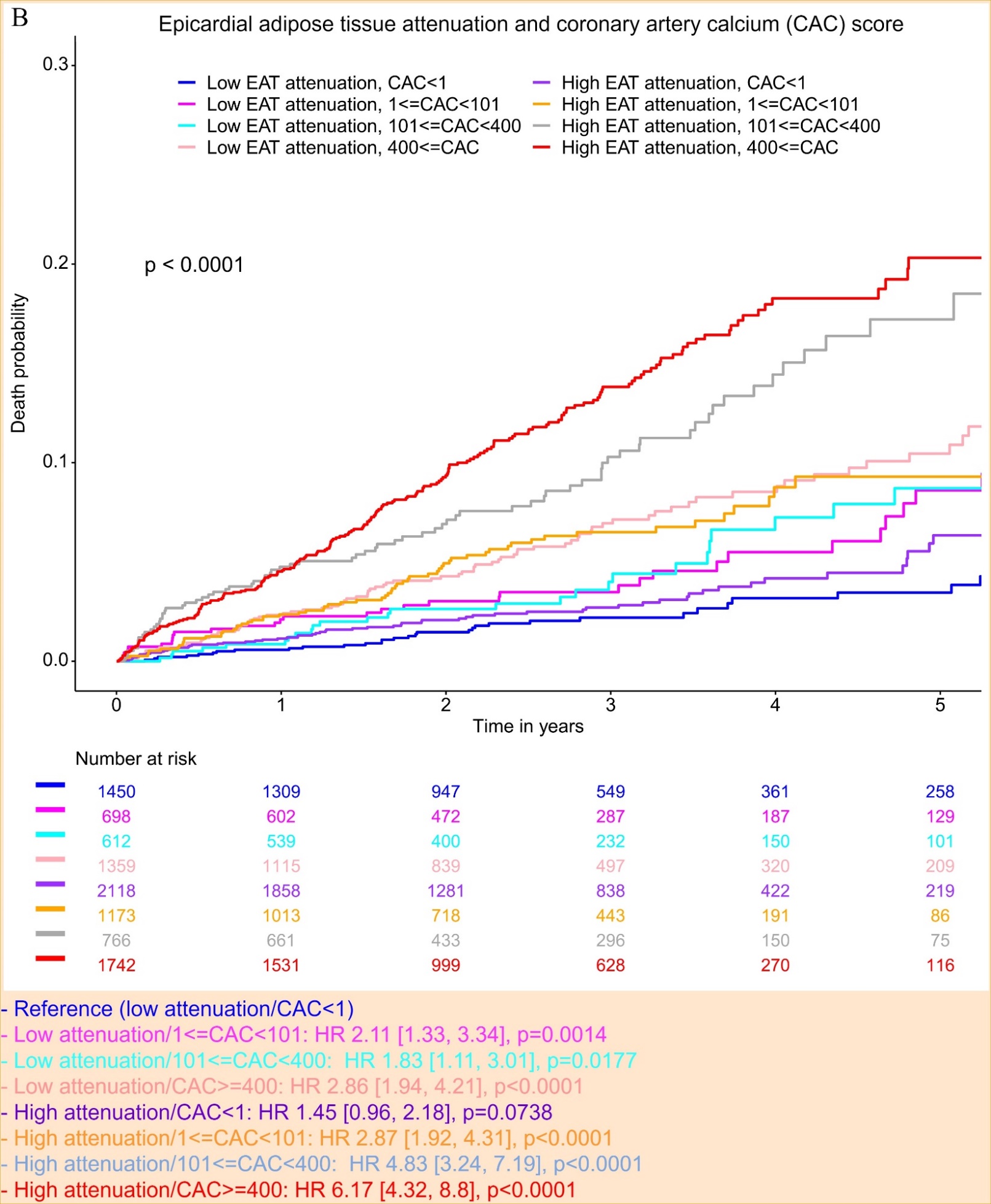

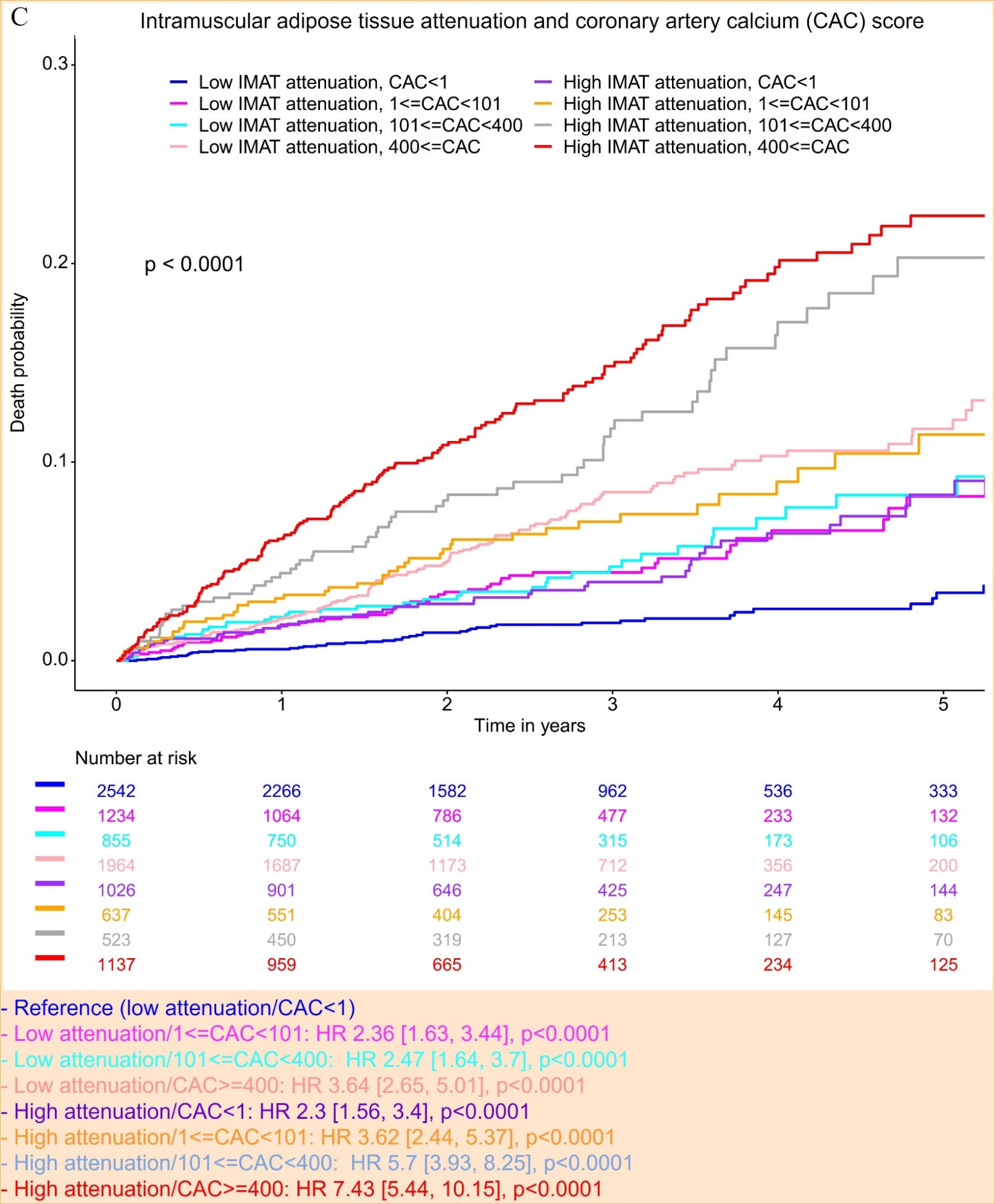

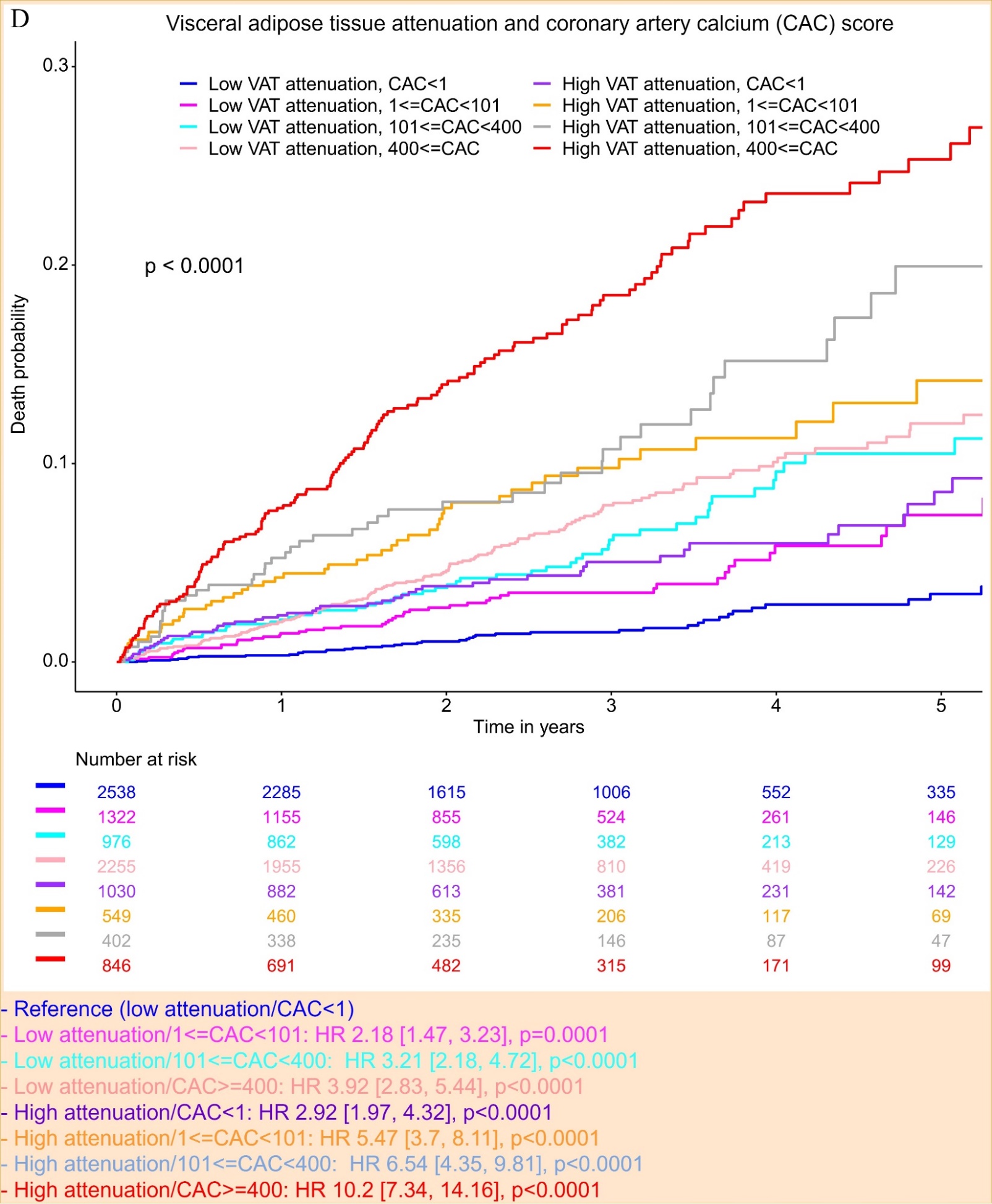

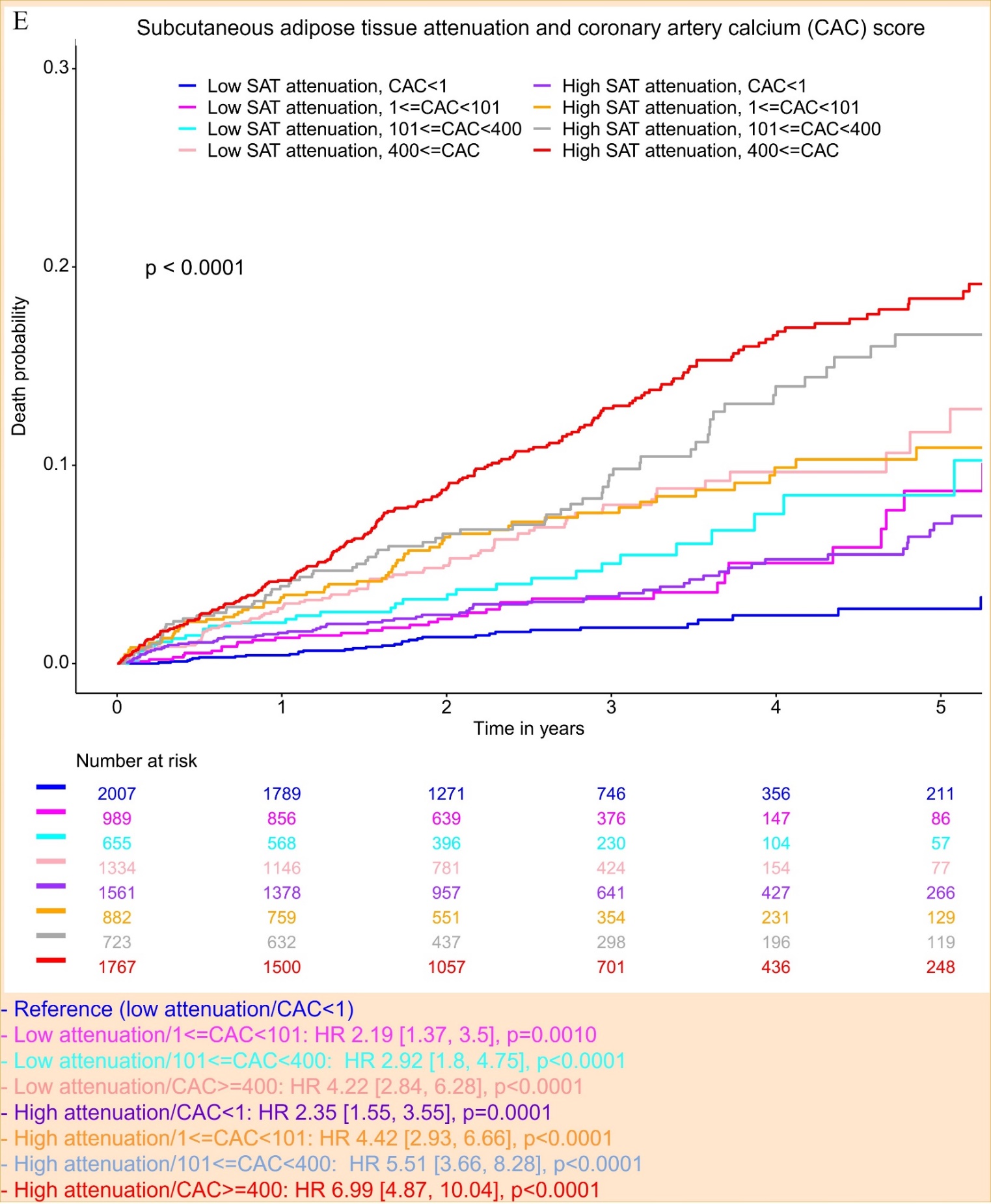

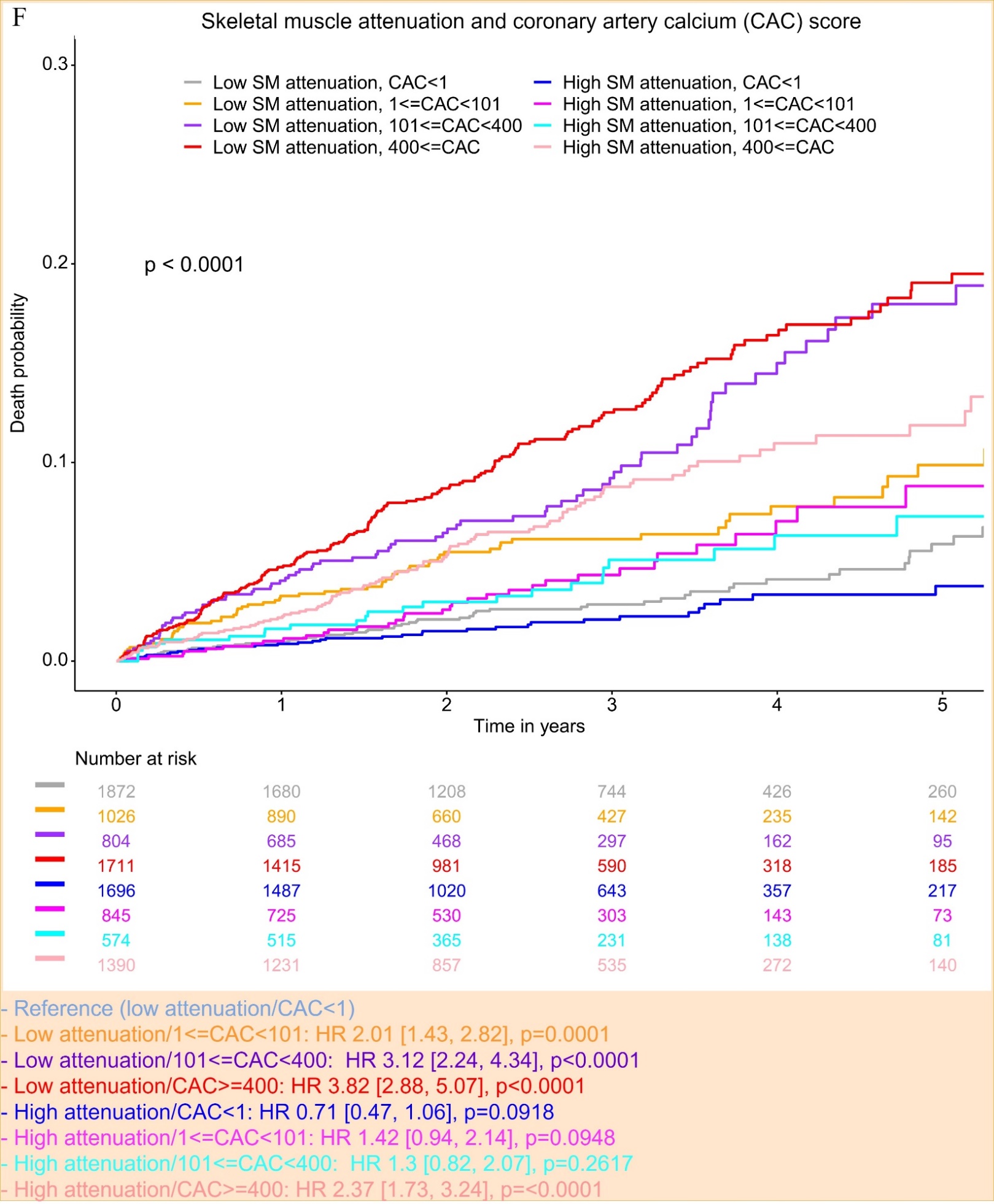

**Supplementary Figure 5.** **Kaplan-Meier curves for risk stratification with volumetric body composition attenuation and coronary artery calcium (CAC) score in all patients.** **A:** bone, **B:** EAT – epicardial adipose tissue, **C:** IMAT – intramuscular adipose tissue, **D:** VAT – visceral adipose tissue, **E:** SAT – subcutaneous adipose tissue, **F:** SM – skeletal muscle. The cutoffs were from Suppl. Table 5. HR – hazard ratio.
